# A stochastic model for Lassa fever infection

**DOI:** 10.64898/2025.12.17.25342531

**Authors:** Praise-God Madueme, Faraimunashe Chirove

**Affiliations:** Department of Mathematics and Applied Mathematics, University of Johannesburg, Auckland Park, 2006, Johannesburg, South Africa

**Keywords:** Extinction, probability, persistence, branching process

## Abstract

This paper looked at the exploration of Lassa fever transmission dynamics through stochastic models which yielded valuable insights into the interplay of factors influencing the probability of extinction and persistence of the virus within a population. By embracing the inherent randomness and variability in the system, the model provided a more realistic representation of the complex ecological and epidemiological dynamics of Lassa fever. We developed the deterministic model using a system of ordinary differential equations and the stochastic model using the Continuous Time Markov Chain. The probability of extinction and persistence underscored the need for a proactive and flexible approach to public health management. Our study revealed that introducing Lassa virus at the onset of an epidemic through various routes affects the likelihood of pathogen extinction. The presence of multiple infection routes increased the probability of pathogen persistence, highlighting complex transmission dynamics. Variations in contact rates, particularly between susceptible rodents and the environment community pathogen load, play a crucial role in influencing pathogen dynamics. This interconnected nature of transmission pathways underscores the factors governing Lassa virus persistence or extinction in a population, providing valuable insights for targeted management and control strategies for Lassa fever.

## 1. Introduction

Lassa fever, a viral hemorrhagic fever precipitated by the Lassa virus, has captured the concern of the scientific community owing to its ominous potential for widespread outbreaks and its distressing high mortality rates. Consequently, it has emerged as a focal point of considerable attention within the realm of infectious diseases. With its intricate epidemiology, transmission pathways, and propensity for large-scale outbreaks, Lassa fever remains a subject of investigation. Researchers seek to understand it better to find ways to reduce its impact. Both deterministic and stochastic models play a pivotal role in the investigation of Lassa fever dynamics. These mathematical models have proven invaluable in deciphering the interplay of variables that govern the spread of the virus and in forecasting potential scenarios, thus aiding in the development of comprehensive control strategies to combat the disease [1, 2].

Many of the models utilized in the study of Lassa fever dynamics are deterministic in nature, which means they overlook the potential influence of randomness in infection transmission. Deterministic models rely on fixed parameters to depict the spread of a disease and are valuable for estimating the average behavior of the disease within a population. By calculating the basic reproduction number, researchers can gauge the likelihood of an outbreak occurring and the effectiveness of control measures. Stochastic models introduce elements of chance and variability into the modeling process, making them particularly valuable for understanding the impact of random events in small populations and for capturing the effects of unpredictable factors such as individual contact patterns. In the context of endemic infections, stochasticity introduces variation around the endemic level, potentially leading to disease extinction through endemic fade-out, provided that the fluctuations are sufficiently substantial [3]. A striking contrast between deterministic and stochastic epidemic models is their respective convergence behaviors. In stochastic models, the sample paths (stochastic solutions) tend to approach the disease-free equilibrium, whereas in deterministic models, the convergence is directed toward an endemic equilibrium [4]. Stochastic models also offer insights into the potential for disease extinction within a finite timeframe. This enables the calculation of the expected duration until the disease is eradicated [4, 5]. Moreover, stochastic models effectively encompass the intrinsic uncertainty and variability present in real-world epidemics, arising from factors such as demographics and the environment. This becomes relevant when dealing with initial low infection numbers [5]. The objective of this study is to construct and comprehensively analyze a stochastic epidemic model, employing a Continuous-Time Markov Chain (CTMC) framework, to elucidate the transmission dynamics of Lassa fever within human, rodent, and virus populations. We use CTMC models to better capture the continuous nature of LF infection progression.

The remaining sections are as follows: In section 2, we introduce a deterministic model. In section 3, we develop the stochastic counterpart of the deterministic model. In section 4, we analyze the stochastic threshold for disease extinction or invasion, and in section 5, we present numerical simulations to illustrate the results. In section 6, we present the discussion of the outcomes, which includes a comparative analysis of the stochastic and deterministic models.

## 2. The deterministic Lassa Fever model

The deterministic model we use is such that the transmission dynamics of Lassa fever is based on the community pathogen load contribution from human, rodent and virus populations. The total human population is *N*_*H*_ (*t*) and is given as

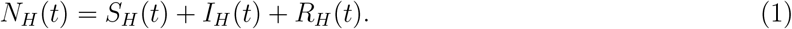

Here, *S*_*H*_ (*t*) is the class of susceptible humans, *I*_*H*_ (*t*) refers to the population of all infected humans, and *R*_*H*_ (*t*) is the class of recovered humans. At any time *t*, the constant recruitment of susceptible humans is given as *π*_1_. All humans die naturally at a rate *µ*_1_ and infected humans die due to the Lassa virus at the rate *δ*. Infected humans recover at the rate *ψ*. Susceptible humans contract Lassa fever through the force of infection

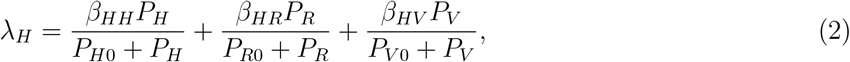

where *β*_*HH*_ is the maximum contact rate of susceptible humans with infected humans, *β*_*HR*_ is the maximum contact rate of susceptible humans with infected rodents, *β*_*HV*_ is the maximum contact rate of susceptible humans with the contaminated environment; *P*_*H*_ is the human community pathogen load (HCPL), *P*_*R*_ is the rodent community pathogen load (RCPL), *P*_*V*_ is the community pathogen load of the virus in the environment (VCPL) and *P*_*H*0_, *P*_*R*0_ and *P*_*V* 0_ are the half-saturation constants of the human, rodent and virus functional response. The HCPL is described as the collection of all individual pathogen loads of humans infected with the Lassa virus in a particular community at a particular time. If one infected human produces *V*_*H*_ virus at the rate *τ*_*H*_, the number of viruses produced by all infected humans is given by *τ*_*H*_ *V*_*H*_ *I*_*H*_ . Here, *V*_*H*_ is the average population of the Lassa virus within a single infected human host. This total infectious reservoir of humans in the community is our *P*_*H*_ . The RCPL is defined as the collection of all individual pathogen loads of rodents infected with the Lassa virus in a particular community at a particular time. If a single infected rodent generates *V*_*R*_ virus at the rate *τ*_*R*_, then *τ*_*R*_*V*_*R*_*I*_*R*_ measures the quantity of Lassa virus generated by all infected rodents. Here, *V*_*R*_ is the average population of the Lassa virus within a single infected rodent host. This total infectious reservoir of rodents in the community is given as *P*_*R*_. The VCPL is the pool that quantifies the total concentration of Lassa virus in the environment at a particular time which is given as *P*_*V*_ .

The total rodent population is *N*_*R*_(*t*) and given as

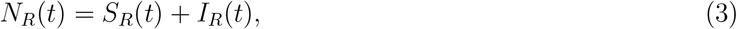

where *S*_*R*_ is the population of rodents susceptible to the virus and *I*_*R*_(*t*) is the population of infected rodents. New recruits move to the rodent population at a constant rate *π*_2_. All rodents die naturally at the rate *µ*_2_ and at a rate *ρ* due to consumption by humans as food. Similarly, susceptible rodents contract the Lassa virus through a force of infection

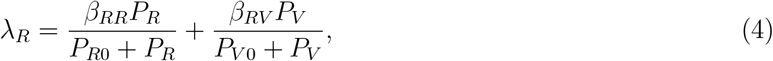

where *β*_*RR*_ is the maximum contact rate of susceptible rodents with infected rodents and *β*_*RV*_ is the maximum contact rate of susceptible rodents with contaminated environment. The rate at which Lassa viral load grows in infected humans is *α*_*H*_ while *α*_*R*_ is the growth rate of Lassa viral load in infected rodents. The classes of infected humans and infected rodents shed the virus in the environment at the rates (1 − *α*_*H*_ ) and (1 − *α*_*R*_) respectively. The natural death/decay rates of the average Lassa viral load within the infected human host, infected rodents, and contaminated environment are *µ*_*PH*_, *µ*_*PR*_, and *µ*_*PV*_ respectively.

The differential equations which describe our model is given by

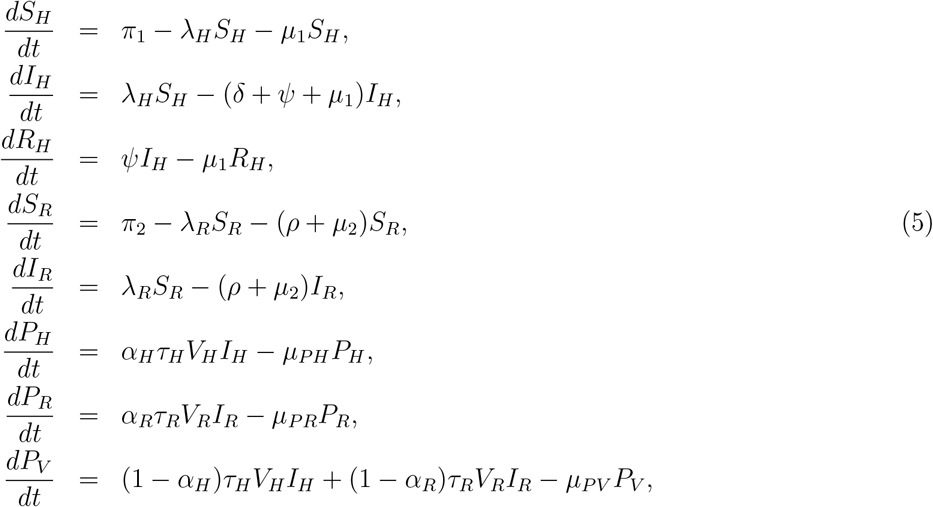

where

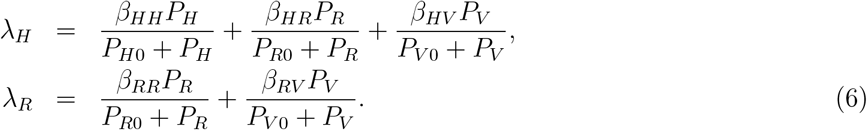

The description of model parameters are shown in table 2.

**Table 1:**
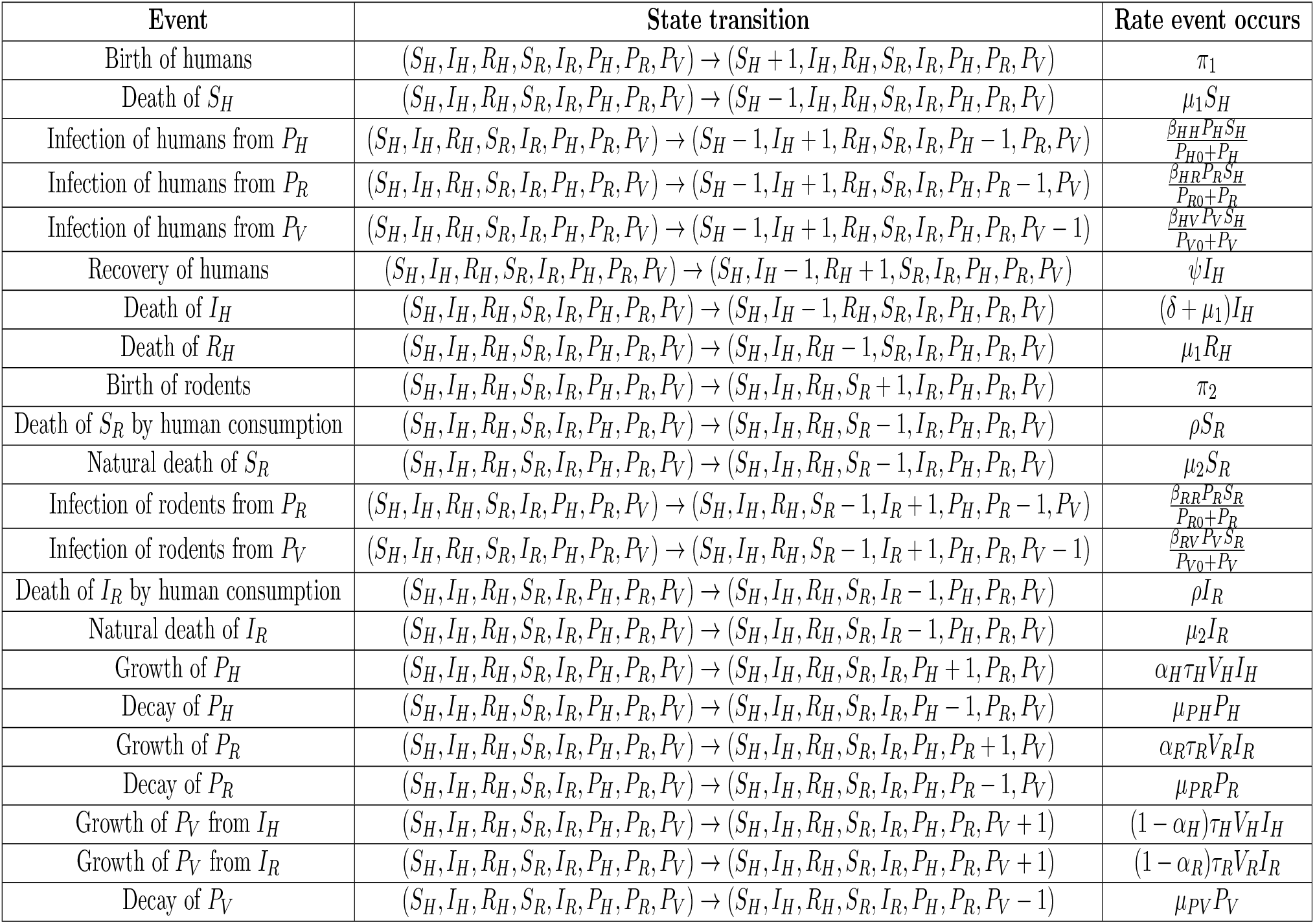
State transitions and rates of occurrence for the CTMC Lassa fever model.

**Table 2:** Parameter values and references.

| Parameters | Meaning | Value | Reference |
| --- | --- | --- | --- |
| $\pi_1$ | Human recruitment rate | 6970.37 | Calculated |
| $\pi_2$ | Rodent recruitment rate | 1692240 | Calculated |
| $\mu_1$ | Human natural death rate | 0.0003488891 | [26] |
| $\mu_2$ | Rodent natural death rate | 0.01923 | [27] |
| $\psi$ | Human recovery rate | 0.0000100436708 | Fitted |
| $\beta_{HH}$ | Contact rate of $S_H$ with $I_H$ | 0.0209 | Fitted |
| $\beta_{HR}$ | Contact rate of $S_H$ with $I_R$ | 0.0122 | Fitted |
| $\beta_{HV}$ | Contact rate of $S_H$ with $P_V$ | 0.0121 | Fitted |
| $\beta_{RR}$ | Contact rate of $S_R$ with $I_R$ | 0.3086 | Fitted |
| $\beta_{RV}$ | Contact rate of $S_R$ with $P_V$ | 0.008 | Fitted |
| $\rho$ | Rodent consumption rate | 0.000085714 | [28, 29] |
| $\delta$ | Human disease-induced death rate | 0.00714285 | [30] |
| $P_{HO}$ | $P_H$ when $\beta_{HH}$ is half (half saturation constant) | 8024.574 | Fitted |
| $P_{RO}$ | $P_R$ when $\beta_{HR}$ is half (half saturation constant) | 5000 | Fitted |
| $P_{VO}$ | $P_V$ when $\beta_{HV}$ is half (half saturation constant) | 4975.426 | Fitted |
| $\alpha_H$ | Growth rate of Lassa viral load in infected humans | 0.5158 | Fitted |
| $\alpha_R$ | Growth rate of Lassa viral load in infected rodents | 0.3086 | Fitted |
| $\tau_H$ | Production rate of $V_H$ virus in one infected human | 0.3086 | Fitted |
| $\tau_R$ | Production rate of $V_R$ virus in one infected rodent | 0.1012 | Fitted |
| $V_H$ | Average LF load within a single infected human host | 502.4574 | Fitted |
| $V_R$ | Average LF load within a single infected rodent host | 50.2457 | Fitted |
| $\mu_{PH}$ | Decay rate of LF load within infected human host | 0.0141 | Fitted |
| $\mu_{PR}$ | Decay rate of LF load within infected rodent host | 0.01899 | Fitted |
| $\mu_{PV}$ | Decay rate of LF load in contaminated environment | 0.008977 | Fitted |

### 2.1. Equilibrium and Stability Analysis

We obtain the equilibrium states of the model by setting the right-hand side of model (5) to zero. At the disease-free equilibrium, there is no Lassa virus and hence no infection in the human and rodent population. The disease-free equilibrium of system (5) is given by

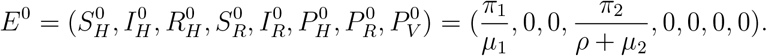

We obtain the basic reproduction number, *R*_0_ of model (5) using the next generation operator approach [6]. Model (5) can be written in the form

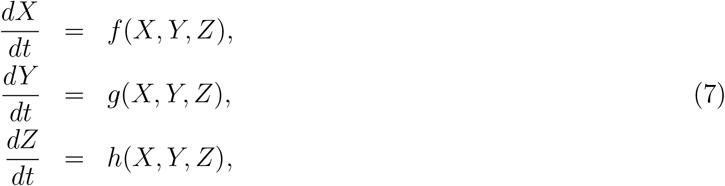

where

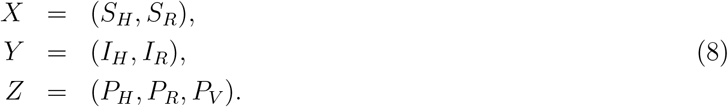

The components of *X* represent the number of susceptibles, the components of *Y* signify the number of infected individuals that do not spread the virus while the components of *Z* signify the number of individuals that can spread the virus. We define

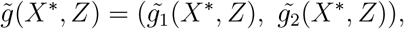

with

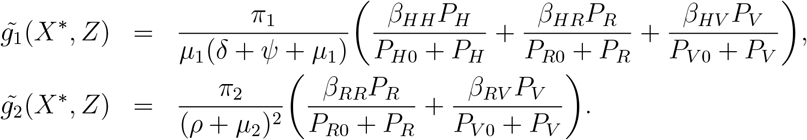

Let 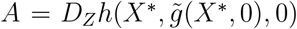 and further assume that *A* can be written as *A* = *M* − *D*, where *M* ≥ 0 and *D* ≥ 0 a diagonal matrix. Then *A* will be

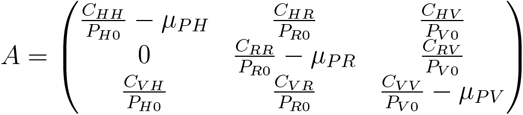

where

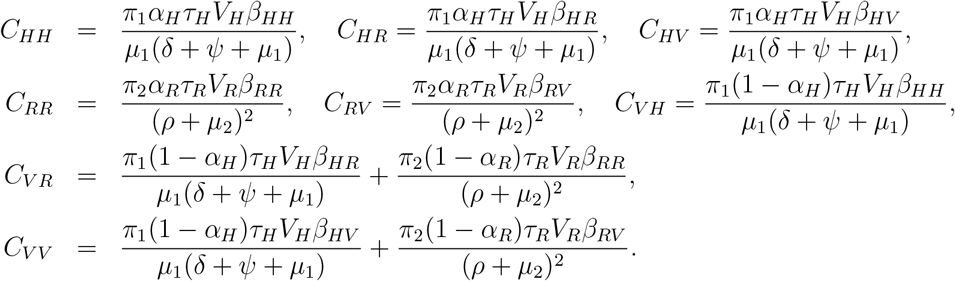

Because *A* = *M* − *D*, we infer that matrices *M* and *D* to be

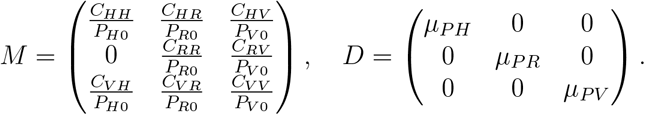

The basic reproduction number is the spectral radius (dominant eigenvalue) of the matrix *MD*^−1^. So

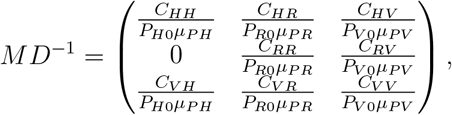

and

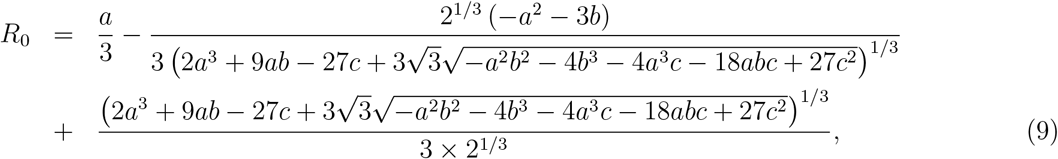

where

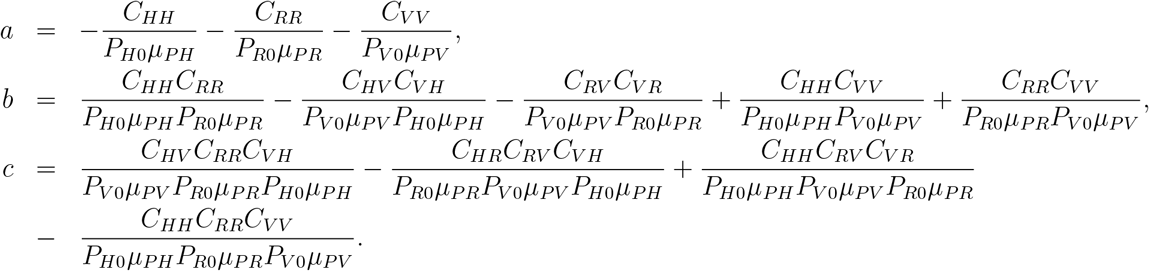

#### Theorem 2.1.1.

*The disease free equilibrium, E*^0^, *for model* (5) *is globally asymptotically stable if R*_0_ ≤ 1.

*Proof*. We apply the approach by Kamgang and Sallet [7] to investigate the global stability of the disease-free equilibrium. It suffices to show that model (5) satisfies hypotheses **H1** to **H5** of the global stability theorem. Let **x**_**1**_ denote the density of the non-infected classes of the model and **x**_**2**_ be the density of the infected classes; that is, **x**_**1**_ = (*S*_*H*_, *R*_*H*_, *S*_*R*_) and **x**_**2**_ = (*I*_*H*_, *I*_*R*_, *P*_*H*_, *P*_*R*_, *P*_*V*_ ). The domain Ω is a compact set and obviously positively invariant.

We consider the sub-system 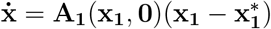:

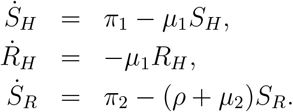

This is a linear system which is globally asymptotically stable at *E*^0^, satisfying the hypotheses **H1** and **H2**.

The matrix **A**_**2**_(**x**) is given by

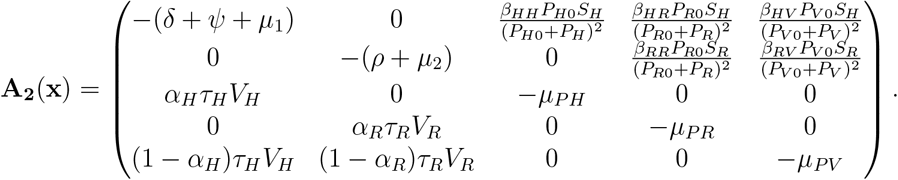

The matrix **A**_**2**_(**x**) is Metzler and irreducible for any **x** ∈ **Ω**, which satisfies hypothesis **H3**.

For hypothesis **H4**, there is a maximum matrix which is uniquely realized in Ω at *E*^0^. This maximum matrix, 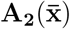, the block of the Jacobian at *E*^0^, is given by

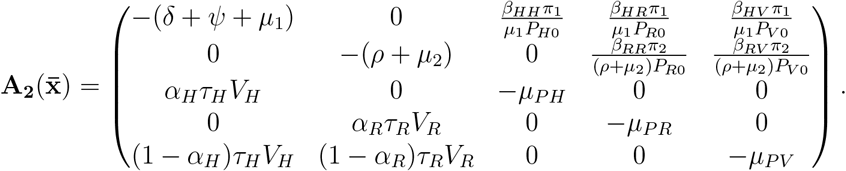

This shows the situation in Corollary 4.4 [7], where the maximum is attained at *E*^0^.

For hypothesis **H5**, we require that 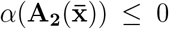 where 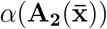 is the stability modulus (spectral bound) of the square matrix 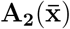. We write 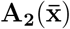 as a block matrix,

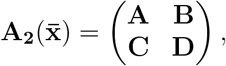

where

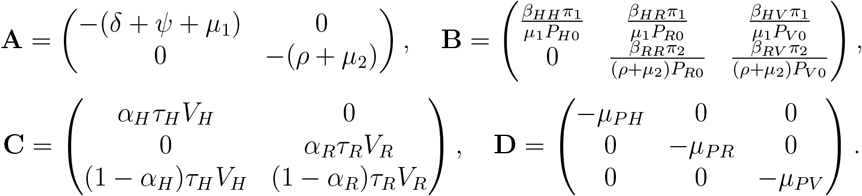

**A** is a Metzler stable matrix and the condition 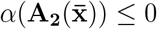 is equivalent to *α*(*D* − *CA*^−1^*B*) ≤ 0. Let

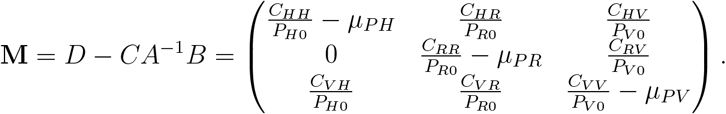

where

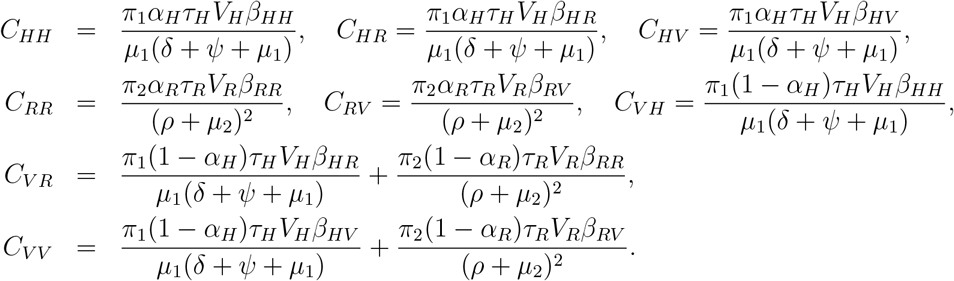

*M* is a stable Metzler matrix if and only if *R*_0_ ≤ 1, that is 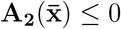 if and only if *R*_0_ ≤ 1. Having satisfied hypothesis **H1** to **H5**, *E*^0^ is globally asymptotically stable if and only if *R*_0_ ≤ 1.

The endemic equilibrium of system (5) is given by

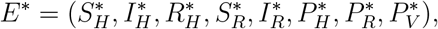

where

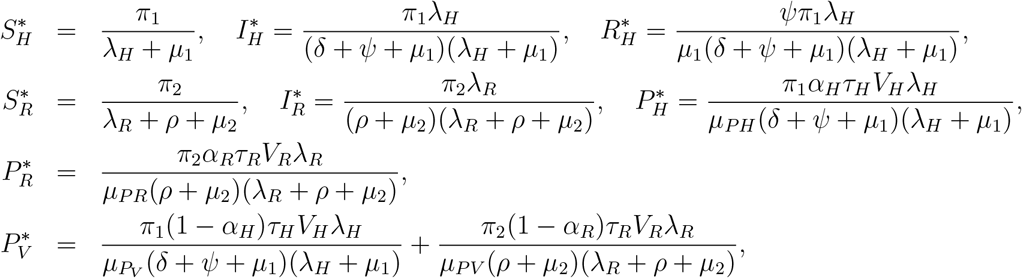

and

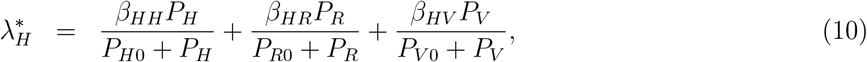

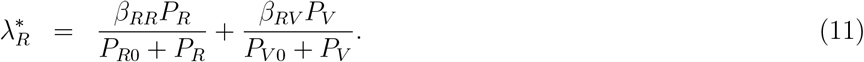

We write equations (10) and (11) as

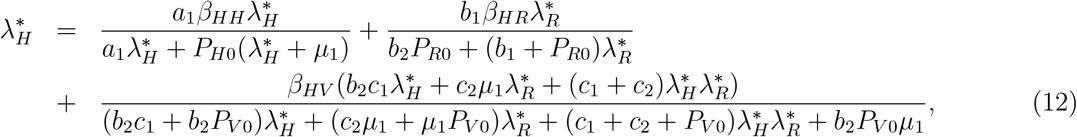

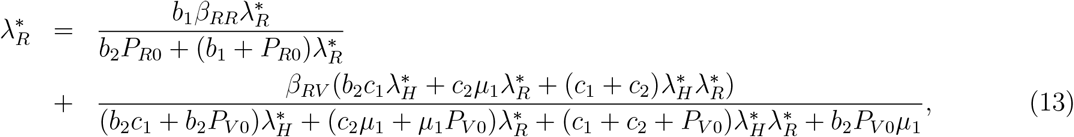

where

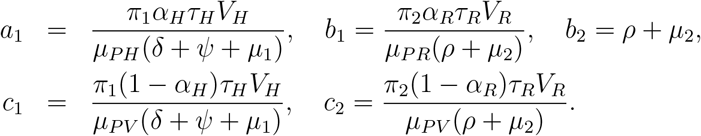

Since the state variables are expressed in terms of 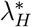 and 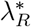, we use the approach in [8, 9] to obtain positive equilibrium points of the model by finding the fixed points of equations (12) and (13) as

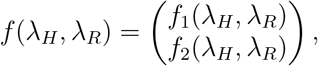

where

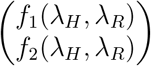

corresponds to the right hand sides of equation (12) and (13).

#### Theorem 2.1.2.

*There exists a unique fixed point* 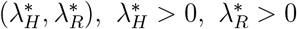 *which satisfies*

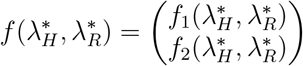

*and corresponds to the endemic equilibrium point E*^*^ *[8, 9]*.

*Proof*. From the first equation, we fix *λ*_*R*_ *>* 0 and look at the real-valued function depending on *λ*_*H*_ :

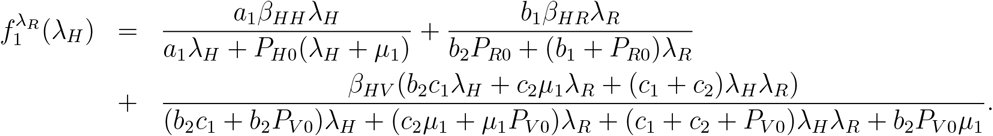

We have that

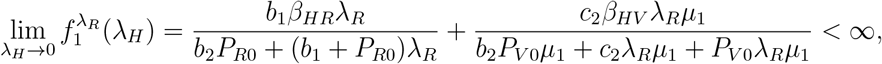

and

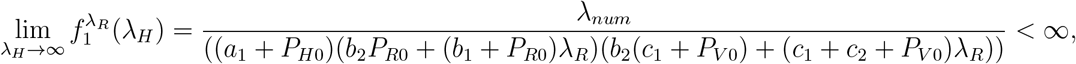

where

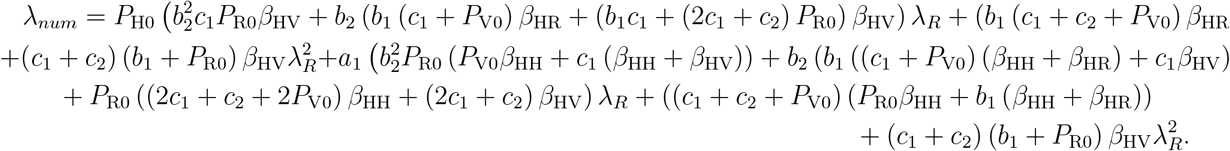

It follows that 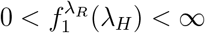 which implies that 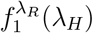 is bounded for every fixed *λ*_*R*_ *>* 0.

Next,

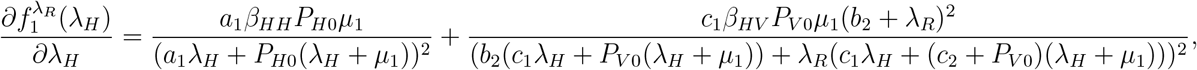

and

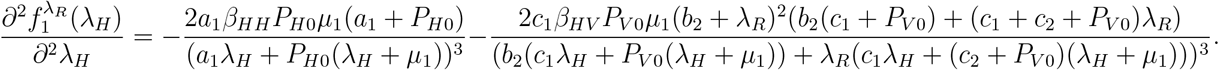

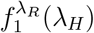 is an increasing concave down function since 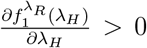 and 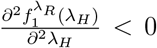. Hence, there is no change in concavity of *f*_1_ in the bounded domain. It follows that there exists a unique 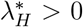 which satisfies 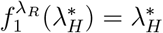.

For this 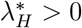, we look at the real-valued function depending on *λ*_*R*_:

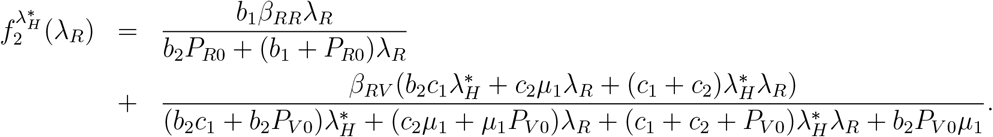

Then,

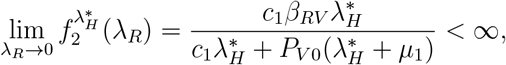

and

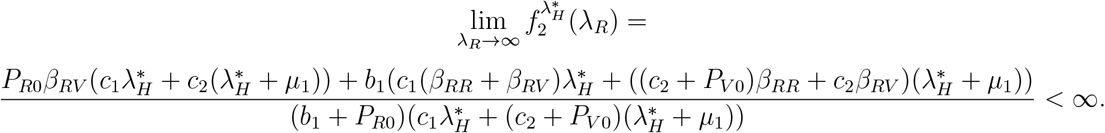

It follows that 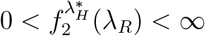 which implies that 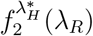 is bounded for every fixed 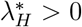.

Next,

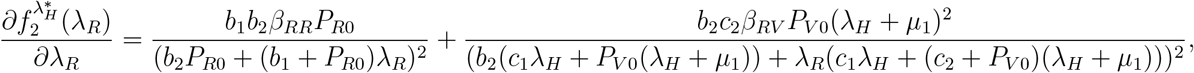

and

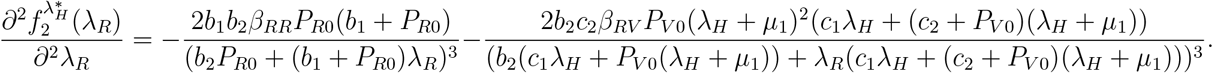

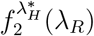 is an increasing concave down function since 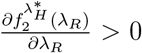 and 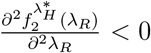. Hence, there is no change in the concavity of *f*_2_ in the positive domain. It follows that there exists a unique 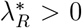 which satisfies 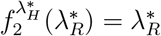.

Therefore, there is a fixed point 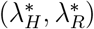 which corresponds to the endemic equilibrium point *E*^*^.

The stability conditions of *E*^*^ are determined by the eigenvalues of the Jacobian matrix of equations (12) and (13) given as

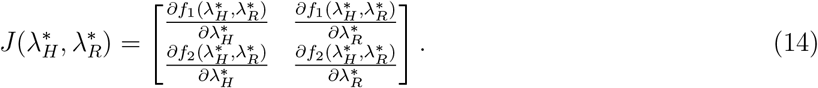

#### Theorem 2.1.3.

*[10] Let λ*_*i*_, *i* = *H, R, be the eigenvalues of the Jacobian matrix* 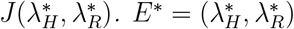 *is asymptotically stable if* |*λ*_*i*_| *<* 1.

*Proof*. Let

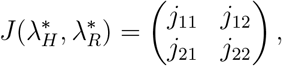

where

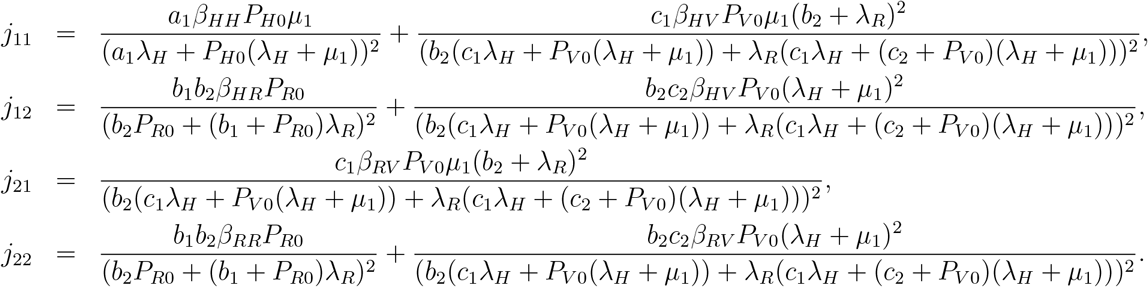

|*λ*_*i*_| *<* 1 corresponds to

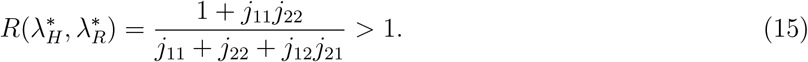

The fixed point 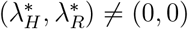 is locally stable when 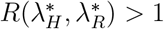.

## 3. The Stochastic Model

Stochastic epidemiological models offer a perspective on disease dynamics by accounting for discrete transitions of individuals between various disease states, in contrast to relying on average transition rates. In stochastic epidemic models, the population’s numbers in each disease category are represented as whole numbers, emphasizing the discrete nature of disease progression [11], and there is consideration that the last infected individual might recover before further transmission occurs, with any subsequent infections requiring reintroduction from external sources [3]. This contrasts with deterministic models, which exhibit a limitation where infections can plummet to extremely low levels, possibly reaching the point of a solitary infected individual, only to resurge at a later time [4]. The introduction of variability in stochastic models can yield dynamics that give other insights that are exhibited by deterministic models [12].

### 3.1. Stochastic model formulation

We develop a stochastic version of the deterministic model (5) by employing a Continuous-Time Markov Chain (CTMC) approach, chosen due to the continuous nature of time and the discrete nature of random variables [4, 13]. This model accounts for the inherent variability in individual birth and death processes, referred to as demographic variability. To facilitate the analysis of the CTMC model, we maintain the same notation for state variables and parameters as that used in the deterministic model (5). In this case, time is considered continuous, denoted as *t* ∈ [0, ∞), while the variables *S*_*H*_, *I*_*H*_, *R*_*H*_, *S*_*R*_, *I*_*R*_, *P*_*H*_, *P*_*R*_, *P*_*V*_ represent discrete random variables for the number of susceptible humans, infected humans, recovered humans, susceptible rodents, infected rodents, human community pathogen load, rodent community pathogen load, and virus community pathogen load, respectively, with finite state spaces,

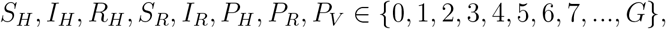

where *G* is a positive integer and represents the maximum size of the population in the finite space [13].

In CTMC models, state transitions can occur at any moment in time, *t*. The transition details and associated rates for the CTMC model are provided in Table 1. The stochastic process

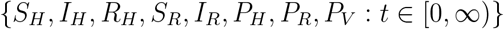

is a multivariate process, and its joint probability function [13] is given by

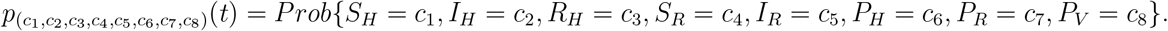

We assume that the stochastic process exhibits temporal homogeneity and adheres to the Markov property. The property dictates that the future state of the process at time (*t* + Δ*t*) solely relies on the current state of the process at time *t*, so that

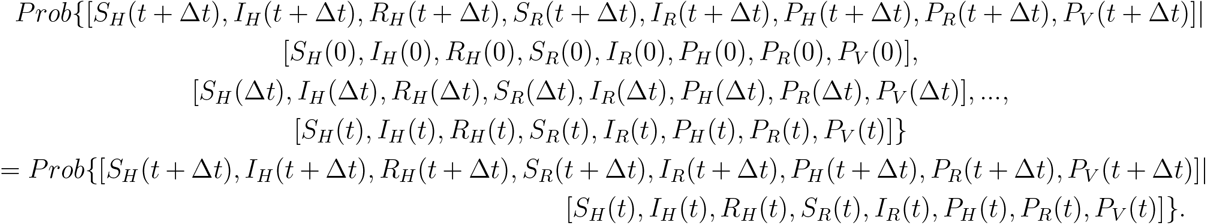

The Markov assumption assures that the time to next event is exponentially distributed [13, 14, 15]. We assume that at most one event occurs during the time interval Δ*t*. The Infinitesimal Transition Probabilities (ITPs) for the stochastic process from state *k* = (*c*_1_, *c*_2_, *c*_3_, *c*_4_, *c*_5_, *c*_6_, *c*_7_, *c*_8_) at time *t* to a new state *k* + *m* = (*d*_1_, *d*_2_, *d*_3_, *d*_4_, *d*_5_, *d*_6_, *d*_7_, *d*_8_) at time (*t* + Δ*t*), is given as

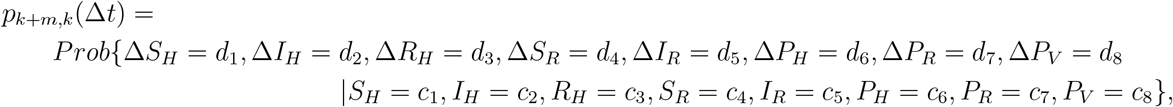

are defined by

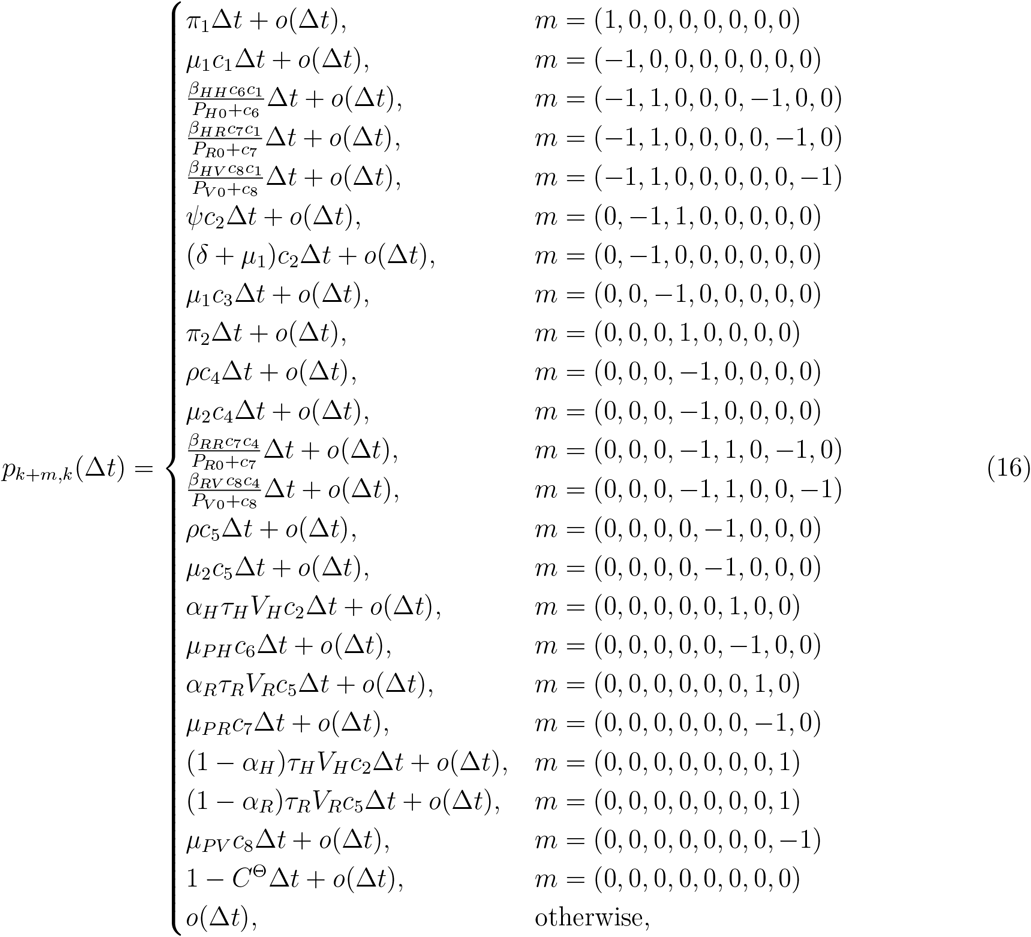

where

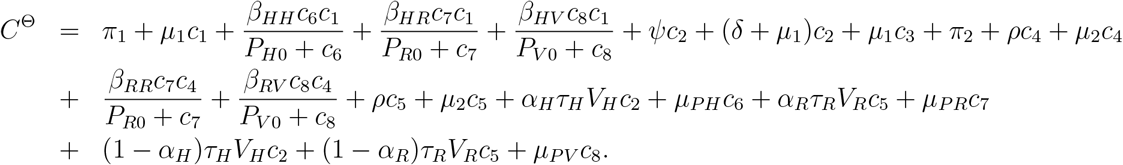

The probability of no change in any of the state variables, *p*(Δ*t*) = 0 is 1 − *C*^Θ^Δ*t* + *o*(Δ*t*). Applying the Markov property to the stochastic process and the ITPs in (16), we can express the state probabilities at time (*t*+Δ*t*) in terms of the state probabilities at time *t* [16]. Thus, at time (*t*+Δ*t*), the state probabilities 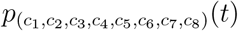 satisfy the following master equation:

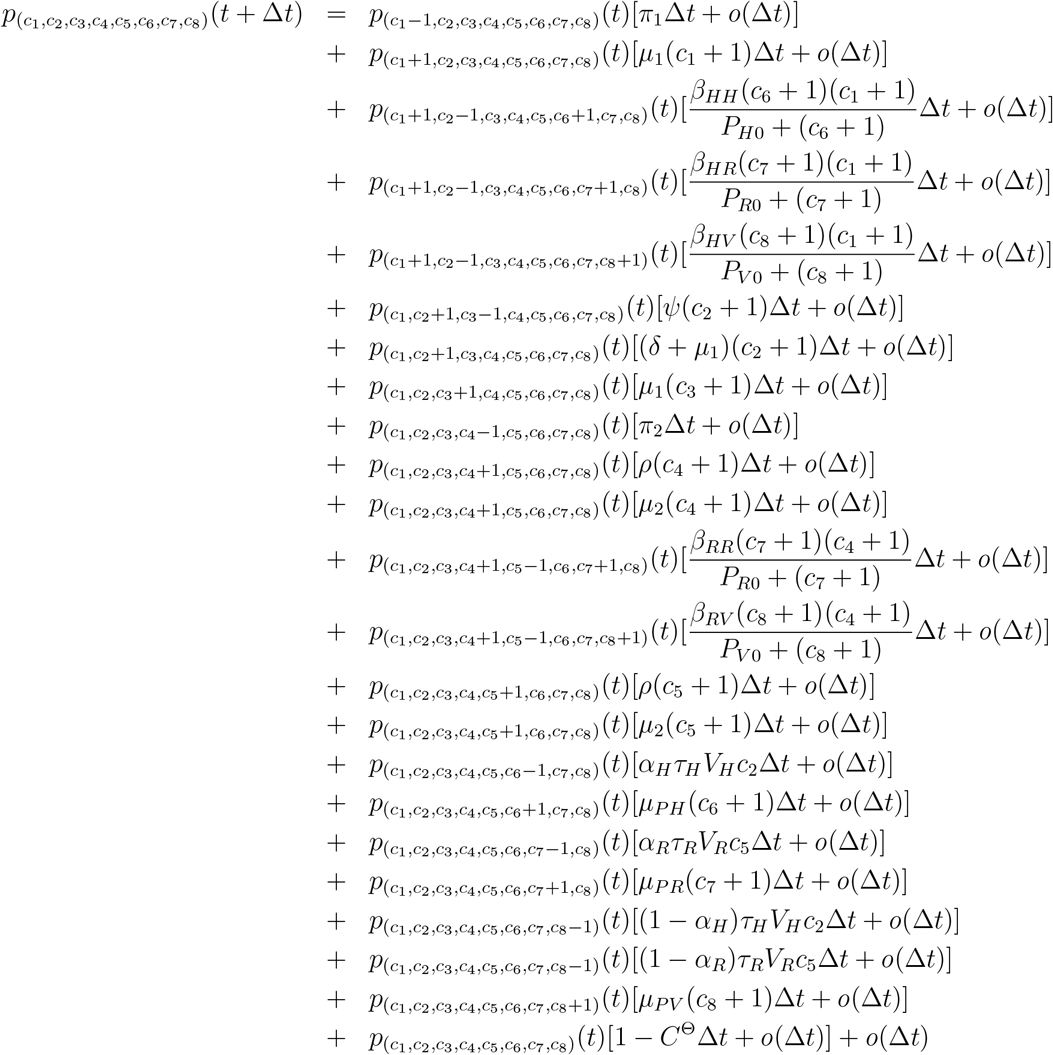

## 4. Stochastic Threshold for Disease Extinction

In stochastic epidemic theory, we can make informed predictions about the occurrence and disappearance of disease outbreaks based on the number of infectious individuals within distinct groups. If a disease emerges from one infectious group with *R*_0_ *>* 1 and if *i* infectives are introduced into a wholly susceptible population, then the probability of a major disease outbreak is approximated by 1(1*/R*_0_)^*i*^ while the probability of disease extinction is approximately (1*/R*_0_)^*i*^ [17]. This principle does not hold true when the infection source involves multiple infectious groups [18]. In scenarios involving multiple infectious groups, stochastic thresholds rely on a combination of two critical elements: the size of each group and the likelihood of disease extinction within each group. It is worth noting that merely having a basic reproduction number *R*_0_ greater than one does not ensure the sustained presence of an infection within a fully susceptible population [3]. Disease extinction is a plausible outcome during the initial phase following the introduction of an infection when the number of infected individuals is still limited. In stochastic models, during this period, the predictions often point to a minor outbreak, in stark contrast to deterministic models, where a major outbreak is always a possibility. Shortly after the introduction of an infection at the early stages of the epidemic process, the depletion of susceptible individuals is minimal. As a result, invasion probabilities can be determined using a linear model that assumes the entire population is susceptible [19, 20]. These invasion probabilities are frequently approximated using the theory of a multitype Galton-Watson Branching Process (GWBP). In GWBP theory, the population is divided into distinct categories, each encompassing a finite number of types characterized by their offspring distribution. The actions of one individual are entirely unrelated to those of others. An individual belonging to a specific type can potentially give rise to offspring of any type, and individuals sharing the same type exhibit an identical distribution of offspring [21, 22].

### 4.1. Galton-Watson Branching Process

The GWBP theory can be applied in the computation of disease invasion and extinction probabilities. The theory is not only relevant to epidemiology but also addresses critical questions concerning extinction and survival in fields like ecology and evolutionary biology, as outlined by Allen [18]. When we possess data regarding the number of infections resulting from a single infectious human or rodent, the GWBP theory becomes a valuable tool for approximating the probabilities of both ultimate disease extinction and the occurrence of a significant disease outbreak. Initially, we provide a comprehensive framework for the branching process, and subsequently, we apply it to our stochastic epidemic model. This application allows us to estimate the likelihood of disease extinction and the probability of a substantial outbreak.

#### Definition 4.1.1.

*[18] A multitype GWBP* 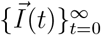 *is a collection of vector random variables* 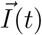, *where each vector consists of k different types*, 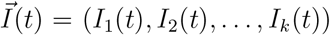 *and each random variable I*_*i*_(*t*) *has k associated offspring random variables for the number of offsprings of type j* = 1, 2, …, *k from a parent of type i*.

The GWBP theory is specifically designed for infectious populations and operates under the assumption that susceptible populations are at the disease-free equilibrium (DFE) [23, 24, 14]. This multitype branching process exhibits linearity in the vicinity of the disease-free equilibrium, possesses a time-homogeneous nature, and involves independent events of birth and death/recovery. As a result, we can establish probability generating functions (pgfs) to model the birth and death/recovery processes within the infectious populations. These pgfs are instrumental in approximating the likelihood of disease extinction and the probability of a substantial outbreak [14].

Suppose we consider infectious individuals of a specific type, denoted as *i* with *I*_*i*_, who contribute to disease transmission by giving birth to individuals of another type, denoted as *j* with *I*_*j*_. The number of offspring produced by an individual of type *i* is independent of the number of offspring generated by either type *i* or type *j, j* ≠ *i*. Also, infectious individuals of type *i* have the same offspring pgf. Let 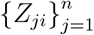 be defined as the offspring random variables for type *i, i* = 1, 2, 3, …, *n* such that *Z*_*ji*_ is the number of offspring of type *j* generated by an infective of type *i*. The offspring pgf for infectious populations *I*_*i*_ is established under the condition where the epidemic process begins with a single infectious individual, i.e., *I*_*i*_(0) = 1, while all other types remain at zero, that is *I*_*j*_(0) = 0, *j* ≠ *i*.

Thus, the offspring pgf *f*_*i*_ : [0, 1]^*n*^ → [0, 1], for type *i* given *I*_*i*_(0) = 1 and *I*_*j*_(0) = 0, *j* ≠ *i*, is defined as

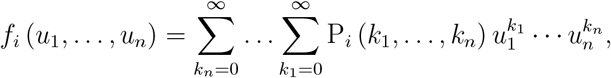

where P_*i*_ (*k*_1_, …, *k*_*n*_) = Prob {*Z*_1*i*_ = *k*_1_, …, *Z*_*ni*_ = *k*_*n*_} is the probability that one infected individual of type *i* gives birth to *k*_*j*_ individuals of type *j* and there is always a fixed point at *f*_*i*_(1, 1, …, 1) = 1 [24, 14, 25]. *f*_*i*_(0, 0, …, 0) denotes the probability of extinction for *I*_*i*_ given that *I*_*i*_(0) = 1 and *I*_*j*_(0) = 0, *i* ≠ *j*.

We introduce the expectation matrix ***M*** = [*m*_*ji*_] as an *n × n*, nonnegative and irreducible matrix where the entry *m*_*ji*_ is the expected number of offsprings of individuals of type *j* produced by an infective individual of type *i*. The elements of matrix ***M*** are calculated from Eq. (14) by differentiating *f*_*i*_ with respect to *u*_*j*_ and then evaluating all the ***u*** variables at 1, that is,

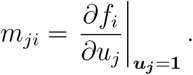

The probability of disease extinction or persistence for the multitype GWBP hinges on the size of the spectral radius of expectation matrix ***M***, *ρ*(***M*** ). If *ρ*(***M***) ≤ 1, then the probability of ultimate disease extinction is one as *t* → ∞ but if *ρ*(***M*** ) *>* 1, then there is a positive probability that the disease may persist [21, 22]. The conditions that determine whether a disease will vanish or persist are presented in the following theorem.

#### Theorem 4.1.1.

*[14, 23, 24, 25] Let the initial sizes for each type be I*_*i*_(0) = *i*_*i*_, *i* = 1, 2, …, *k. Suppose the generating functions f*_*i*_ *for each of the k types are nonlinear functions of u*_*j*_ *with some f*_*i*_(0, 0, …, 0) *>* 0. *Also, suppose that the expectation matrix* ***M*** = [*m*_*ji*_] *is an n × n nonnegative and irreducible matrix, and ρ*(***M*** ) *is the spectral radius of matrix M*.

i. *If ρ*(***M*** ) *<* 1 *or ρ*(***M*** ) = 1 *(subcritical and critical case respectively), then the probability of ultimate extinction is one:*

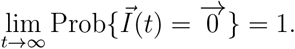
ii. *If ρ*(***M*** ) *>* 1 *(supercritical case), then the probability of ultimate disease extinction is less than one:*

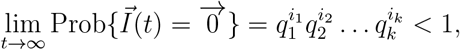

*where* (*q*_1_, *q*_2_, …, *q*_*k*_) *is the unique fixed point of the k offspring pgf, f*_*i*_ (*q*_1_, *q*_2_, …, *q*_*k*_) = *q*_*i*_ *and* 0 *< q*_*i*_ *<* 1, *i* = 1, 2, …, *k. The value of q*_*i*_ *is the probability of disease extinction for infectives of type i and the probability of an outbreak is approximately*

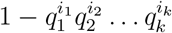

#### 4.1.1. Application of the Multitype GWBP

We apply the branching process to the CTMC model by defining offspring pgfs of all infected classes in the model. We assume that the susceptible human and rodent populations are sufficiently large and are at the DFE. The offspring pgf for *I*_*H*_ (0) = 1, *I*_*R*_(0) = 0, *P*_*H*_ (0) = 0, *P*_*R*_(0) = 0, *P*_*V*_ (0) = 0 is given by

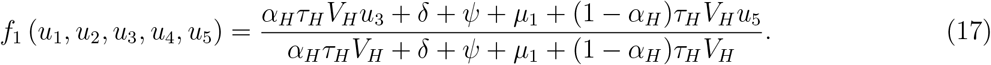

Epidemiologically, we can interpret the terms as follows: *α*_*H*_ *τ*_*H*_ *V*_*H*_ */*(*α*_*H*_ *τ*_*H*_ *V*_*H*_ + *δ* + *ψ* + *µ*_1_ + (1 − *α*_*H*_ )*τ*_*H*_ *V*_*H*_ ) shows the probability that an infected human interacts with a susceptible human to produce the human pathogen load. The term (1 − *α*_*H*_ )*τ*_*H*_ *V*_*H*_ */*(*α*_*H*_ *τ*_*H*_ *V*_*H*_ + *δ* + *ψ* + *µ*_1_ + (1 − *α*_*H*_ )*τ*_*H*_ *V*_*H*_ ) shows the probability that an infected human interacts with a susceptible human to also give birth to the virus pathogen load. The term *δ* +*ψ* +*µ*_1_*/*(*α*_*H*_ *τ*_*H*_ *V*_*H*_ +*δ* +*ψ* +*µ*_1_ +(1 − *α*_*H*_ )*τ*_*H*_ *V*_*H*_ ) shows the probability that an infected human dies before transmitting the virus which results in zero infectious humans.

The offspring pgf for *I*_*H*_ (0) = 0, *I*_*R*_(0) = 1, *P*_*H*_ (0) = 0, *P*_*R*_(0) = 0, *P*_*V*_ (0) = 0 is given by

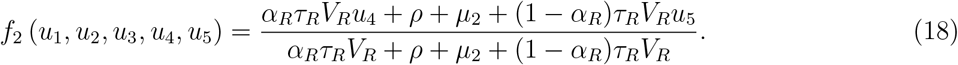

We interpret the terms epidemiologically as follows: the term *α*_*R*_*τ*_*R*_*V*_*R*_*/α*_*R*_*τ*_*R*_*V*_*R*_ + *ρ* + *µ*_2_ + (1 − *α*_*R*_)*τ*_*R*_*V*_*R*_ shows the probability that an infected rodent interacts with a susceptible rodent to produce the rodent pathogen load. The term (1 − *α*_*R*_)*τ*_*R*_*V*_*R*_*/α*_*R*_*τ*_*R*_*V*_*R*_ + *ρ* + *µ*_2_ + (1 − *α*_*R*_)*τ*_*R*_*V*_*R*_ shows the probability that an infected rodent interacts with a susceptible rodent to also give birth to the virus pathogen load. The term *ρ* + *µ*_2_*/α*_*R*_*τ*_*R*_*V*_*R*_ + *ρ* + *µ*_2_ + (1 − *α*_*R*_)*τ*_*R*_*V*_*R*_ shows the probability that an infected rodent dies before transmitting the virus which results in zero infections rodents.

The offspring pgf for *I*_*H*_ (0) = 0, *I*_*R*_(0) = 0, *P*_*H*_ (0) = 1, *P*_*R*_(0) = 0, *P*_*V*_ (0) = 0 is given by

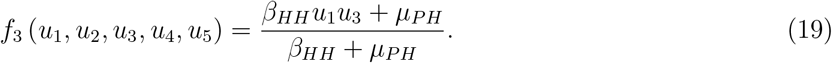

We interpret the terms as follows: the term *β*_*HH*_ */β*_*HH*_ +*µ*_*PH*_ shows the probability that human community pathogen load interacts with a susceptible human to produce an infected human and also give birth to the human community pathogen load. The term *µ*_*PH*_ */β*_*HH*_ + *µ*_*PH*_ shows the probability that the human community pathogen load dies before transmitting the virus resulting in zero human infections.

The offspring pgf for *I*_*H*_ (0) = 0, *I*_*R*_(0) = 0, *P*_*H*_ (0) = 0, *P*_*R*_(0) = 1, *P*_*V*_ (0) = 0 is given by

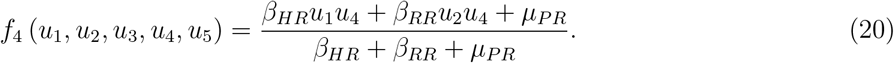

We interpret the terms as follows: the term *β*_*HR*_*/β*_*HR*_ + *β*_*RR*_ + *µ*_*PR*_ shows the probability that rodent community pathogen load interacts with a susceptible human to produce an infected human and also give birth to rodent community pathogen load. The term *β*_*RR*_*/β*_*HR*_ + *β*_*RR*_ + *µ*_*PR*_ shows the probability that rodent community pathogen load interacts with a susceptible rodent to produce an infected rodent and also give birth to rodent community pathogen load. The term *µ*_*PR*_*/β*_*HR*_ +*β*_*RR*_ +*µ*_*PR*_ shows the probability that the rodent community pathogen load dies before transmitting the virus resulting in zero human and rodent infections.

The offspring pgf for *I*_*H*_ (0) = 0, *I*_*R*_(0) = 0, *P*_*H*_ (0) = 0, *P*_*R*_(0) = 0, *P*_*V*_ (0) = 1 is given by

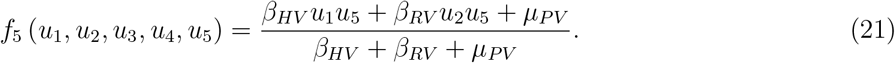

We interpret the terms as follows: the term *β*_*HV*_ */β*_*HV*_ + *β*_*RV*_ + *µ*_*PV*_ shows the probability that the virus community pathogen load interacts with a susceptible human to produce an infected human and also give birth to virus community pathogen load. The term *β*_*RV*_ */β*_*HV*_ + *β*_*RV*_ + *µ*_*PV*_ shows the probability that the virus community pathogen load interacts with a susceptible rodent to produce an infected rodent and also give birth to virus community pathogen load. The term *µ*_*PV*_ */β*_*HV*_ + *β*_*RV*_ + *µ*_*PV*_ shows the probability that the virus community pathogen load dies before transmitting the virus resulting in zero human, rodent and virus infections.

The elements *m*_*ji*_ of the expectation matrix obtained from the offspring pgfs (17) to (21) are given by

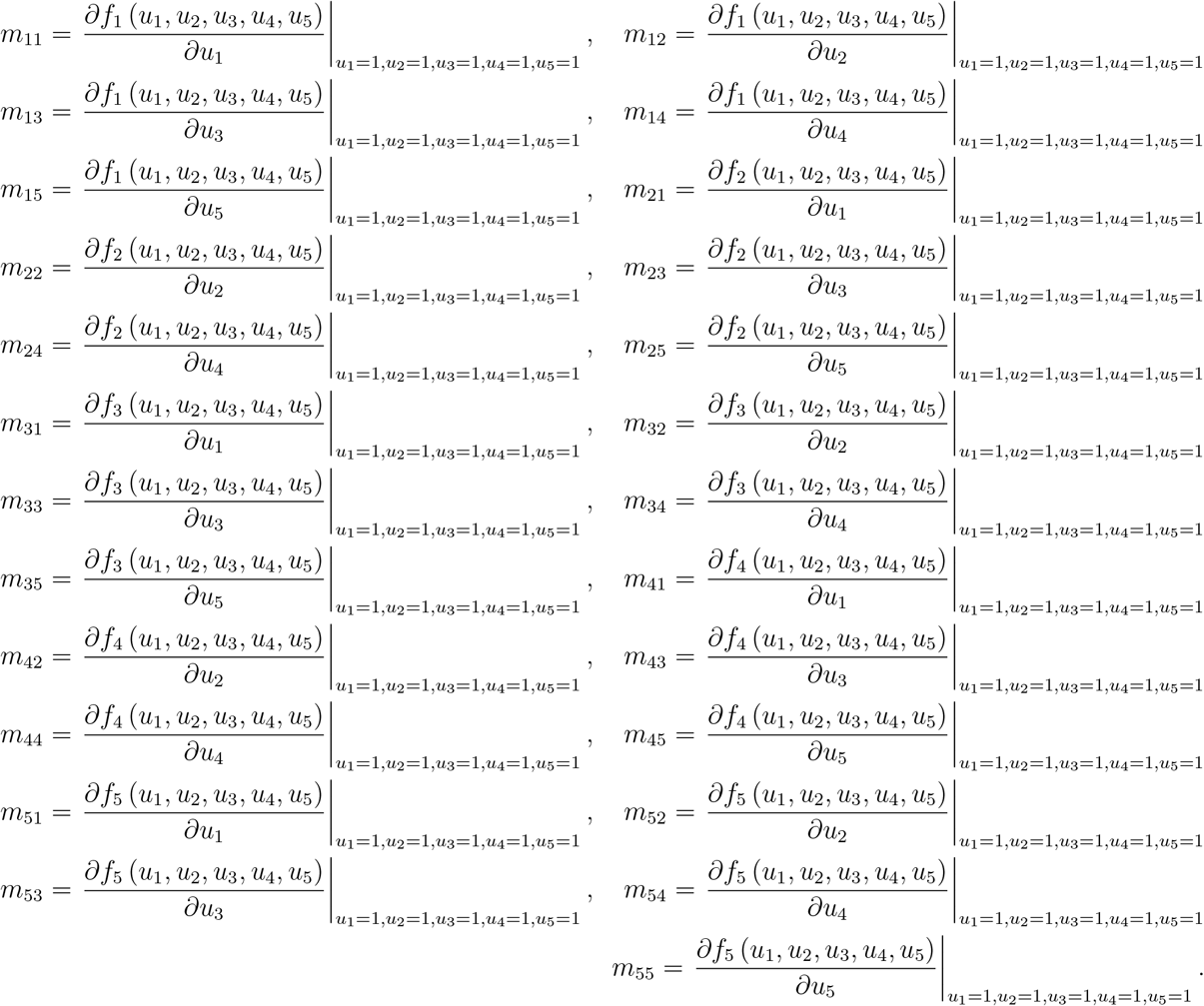

This leads to the expectation matrix

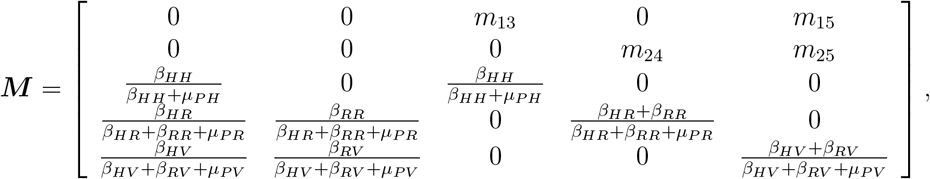

where

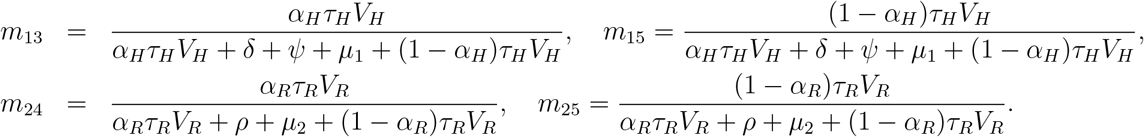

Because of the complex nature of the problem, we use the numerical values of the parameters in table 2 to find the characteristic equation

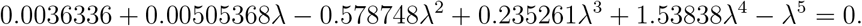

This leads to *ρ*(***M***) = 1.41957 *>* 1. The quantity *ρ*(***M***) is a stochastic threshold for disease extinction or persistence in the human, rodent and virus population. The probability of disease extinction is 1 if *ρ*(***M***) ≤ 1 which leads to the elimination of the disease from the population. There is positive probability of disease persistence if *ρ*(***M***) *>* 1. The function of the *ρ*(***M***) in stochastic epidemic models is similar to that of *R*_0_ in deterministic models. Allen and van den Driessche [23] established the general relationship that exists between extinction threshold *ρ*(*M*) and ℛ_0_ in stochastic and deterministic epidemic models given by

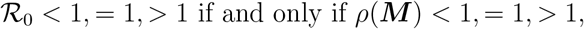

and we can easily show that our model satisfies this relationship. For the supercritical case, *ρ*(***M*** ) *>* 1 (and ℛ_0_ *>* 1 ), there is a positive probability of a major disease outbreak occurring. This implies that there exists a fixed point of the offspring pgfs on (0, 1)^5^ which gives the probability of disease extinction of which one minus this probability is the probability of a major outbreak [14]. The fixed point can be found by setting *f*_*i*_ (*q*_1_, *q*_2_, *q*_3_, *q*_4_, *q*_5_) = *q*_*i*_, *q*_*i*_, *q*_*i*_, *q*_*i*_, *q*_*i*_ ∈ (0, 1), *i* = 1, 2, 3, 4, 5. From the pgfs (17) to (21), we compute the fixed point by simultaneously solving the equations

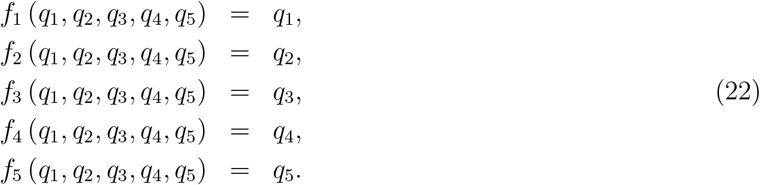

The values *q*_1_, *q*_2_, *q*_3_, *q*_4_, *q*_5_ are the probabilities of ultimate disease extinction infected humans, infected rodents, human community pathogen load, rodent community pathogen load and virus community pathogen load respectively. The quintet (*q*_1_, *q*_2_, *q*_3_, *q*_4_, *q*_5_) = (1, 1, 1, 1, 1) is always a solution but there may exist another fixed point [24]. Thus, we solve the following system of equations:

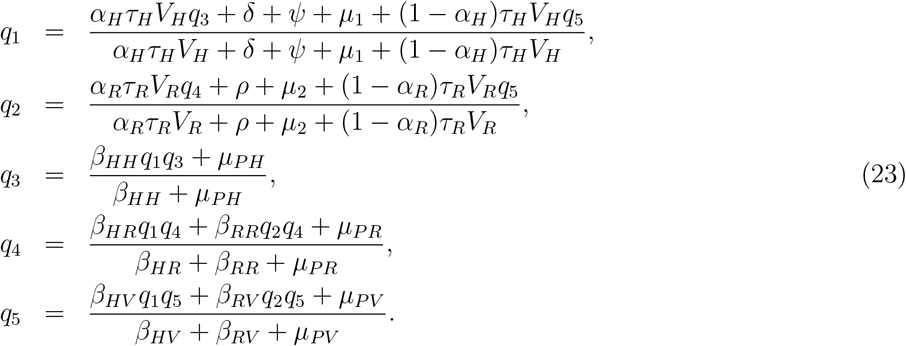

It is difficult to find the 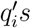 in closed form so we input the numerical values of the parameters from table 2 to get

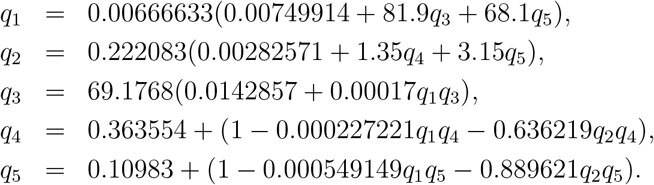

We solve to get

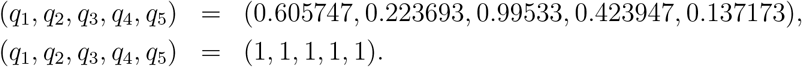

Epidemiologically, *q*_1_ means that an infectious human will successfully transmit the virus to a susceptible human, producing the human pathogen load or gives birth to the virus pathogen load or die before transmitting the virus with probability 0.605747; The term *q*_2_ means that an infectious rodent will successfully transmit the virus to a susceptible rodent, producing the rodent pathogen load or gives birth to the virus pathogen load or die before transmitting the virus with probability 0.223693; The term *q*_3_ means that the interaction between susceptible human and the human community pathogen load gives birth to both infected humans and increases the human community pathogen load or die before transmitting the virus with probability 0.99533; The term *q*_4_ shows that the interaction between susceptible humans, susceptible rodents and the rodent community pathogen load gives birth to both infected humans and infected rodents, and increases the rodent community pathogen load or die before transmitting the virus with probability 0.423947; and the term *q*_5_ shows that the interaction between susceptible humans, susceptible rodents and the virus community pathogen load gives birth to both infected humans and infected rodents, and increases the rodent community pathogen load or die before transmitting the virus with probability 0.137173.

We compute the probability of disease extinction and of an outbreak using *q*_1_, *q*_2_, *q*_3_, *q*_4_, *q*_5_. If *I*_*H*_ (0) = *x*_1_, *I*_*R*_(0) = *x*_2_, *P*_*H*_ (0) = *x*_3_, *P*_*R*_(0) = *x*_4_, *P*_*V*_ (0) = *x*_5_ are the initial sizes of infected humans, infected rodents, human community pathogen load, rodent community pathogen load, and virus community pathogen load, respectively, then the probability of ultimate disease extinction is approximately

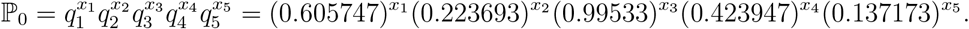

Hence, the probability of a major disease outbreak ℙ_*m*_ is given by

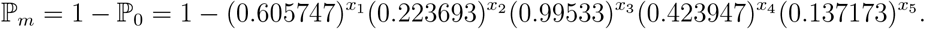

## 5. Numerical Simulations

In this section, we illustrate the dynamics of Lassa fever transmission for the ODE and CTMC models, and approximate the probability of disease extinction or persistence. Further, we vary community pathogen load contact rate parameter values in order to establish their impact on the probability of disease extinction. The final time used in all the simulations ranges from 0 to 3000 weeks. The value of the reproduction number is 1.428.

### 5.1. Probability of disease extinction or persistence

The probability of disease extinction ℙ_0_, for different initial values is calculated from the multitype branching process and is in table 3. The fixed point of the offspring pgfs in (0, 1)^5^ is

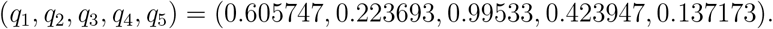

**Table 3:** The probability extinction calculated from branching process. The probabilities are calculated with a stable endemic equilibrium, *E*^*^. Parameter values are as in table 2 with initial conditions *S*_*H*_ (0) = 1997.8662, *I*_*H*_ (0) = 1, *R*_*H*_ (0) = 0, *S*_*R*_(0) = 1000, *I*_*R*_(0) = 1, *P*_*H*_ (0) = 1, *P*_*R*_(0) = 1, *P*_*V*_ (0) = 1.

| $x_1$ | $x_2$ | $x_3$ | $x_4$ | $x_5$ | $\mathbb{P}_0$ |
| --- | --- | --- | --- | --- | --- |
| 1 | 0 | 0 | 0 | 0 | 0.605747 |
| 0 | 1 | 0 | 0 | 0 | 0.223693 |
| 0 | 0 | 1 | 0 | 0 | 0.99533 |
| 0 | 0 | 0 | 1 | 0 | 0.423947 |
| 0 | 0 | 0 | 0 | 1 | 0.137173 |
| 1 | 1 | 0 | 0 | 0 | 0.135501364 |
| 1 | 0 | 1 | 0 | 0 | 0.602918162 |
| 1 | 0 | 0 | 1 | 0 | 0.256804623 |
| 1 | 0 | 0 | 0 | 1 | 0.083092133 |
| 0 | 1 | 1 | 0 | 0 | 0.222648354 |
| 0 | 1 | 0 | 1 | 0 | 0.094833976 |
| 0 | 1 | 0 | 0 | 1 | 0.03068464 |
| 0 | 0 | 1 | 1 | 0 | 0.421967168 |
| 0 | 0 | 1 | 0 | 1 | 0.136532402 |
| 0 | 0 | 0 | 1 | 1 | 0.058154082 |
| 1 | 1 | 1 | 0 | 0 | 0.134868572 |
| 1 | 1 | 0 | 1 | 0 | 0.057445397 |
| 1 | 1 | 0 | 0 | 1 | 0.018587129 |
| 1 | 0 | 1 | 1 | 0 | 0.255605346 |
| 1 | 0 | 1 | 0 | 1 | 0.082704093 |
| 1 | 0 | 0 | 1 | 1 | 0.035226661 |
| 0 | 1 | 1 | 1 | 0 | 0.094391102 |
| 0 | 1 | 1 | 0 | 1 | 0.030541343 |
| 0 | 1 | 0 | 1 | 1 | 0.013008661 |
| 0 | 0 | 1 | 1 | 1 | 0.057882502 |
| 1 | 1 | 1 | 1 | 0 | 0.057177127 |
| 1 | 1 | 1 | 0 | 1 | 0.018500327 |
| 1 | 0 | 1 | 1 | 1 | 0.035062152 |
| 0 | 1 | 1 | 1 | 1 | 0.012947911 |
| 1 | 1 | 1 | 1 | 1 | 0.007843158 |

In Table 3, we observe that the probability of disease extinction varies depending on the initial source of the virus. When the virus is introduced by the HCPL or an infected human at the outset of the epidemic, the likelihood of extinction is high with probabilities of 0.995 and 0.61 respectively. The probabilities are 0.22 and 0.42 if the virus is introduced by an infected rodent or the RCPL, respectively while the introduction of the virus by the VCPL at the beginning of the infection has a extinction probability of 0.14. If there is a single initial contributor to infection, the human contributions and the rodents initiators of infection leads to higher probabilities of disease extinction while disease extinction is least when the initiator is the virus in the environment. In a combination of two initial contributors to infection, *x*_1_ and *x*_3_ give a higher probability of infection extinction and the least comes from *x*_2_ and *x*_5_. In a combination of 3 infected individuals starting off the infection, the highest probability of extinction comes from *x*_1_, *x*_3_ and *x*_4_ and the least from *x*_2_, *x*_4_ and *x*_5_. For combination of 4 infected individuals starting off the infection, highest probability of infection extinction comes from *x*_1_, *x*_2_, *x*_3_ and *x*_4_ and least from *x*_2_, *x*_3_, *x*_4_ and *x*_5_. Examining scenarios where the virus is introduced by all infectious classes in the early stages of the epidemic yield probability of disease extinction that are significantly small. This trend can be associated with the observation that the probability of disease persistence increases with the number of infected individuals across the human, rodent, and virus populations. In essence, a higher load of infected community members contributes to greater odds of disease survival. A combination of virus in the environment in conjunction with all other sources of infection is associated with lower probability of disease extinction. This highlights the threat brought by environmental transmission.

Figure 2 shows a three sample paths simulation showing that the infection persists when infection is initiated by all infectious classes. The sample paths from the CTMC model follow the trajectory of the ODE solutions. Some disease sample paths go to extinction at the outset of the infection and rise after some finite time (see figures 2b and 2g). This suggests a small possibility of disease extinction even when infection is initiated by all infectious classes. We also see from figure 2c that the sample paths are associated with more human recoveries than what is predicted from the deterministic model in a perceived situation of disease persistence. The possibility of more humans recovering suggest possibility of disease extinction over a finite time period.

**Figure 1:**
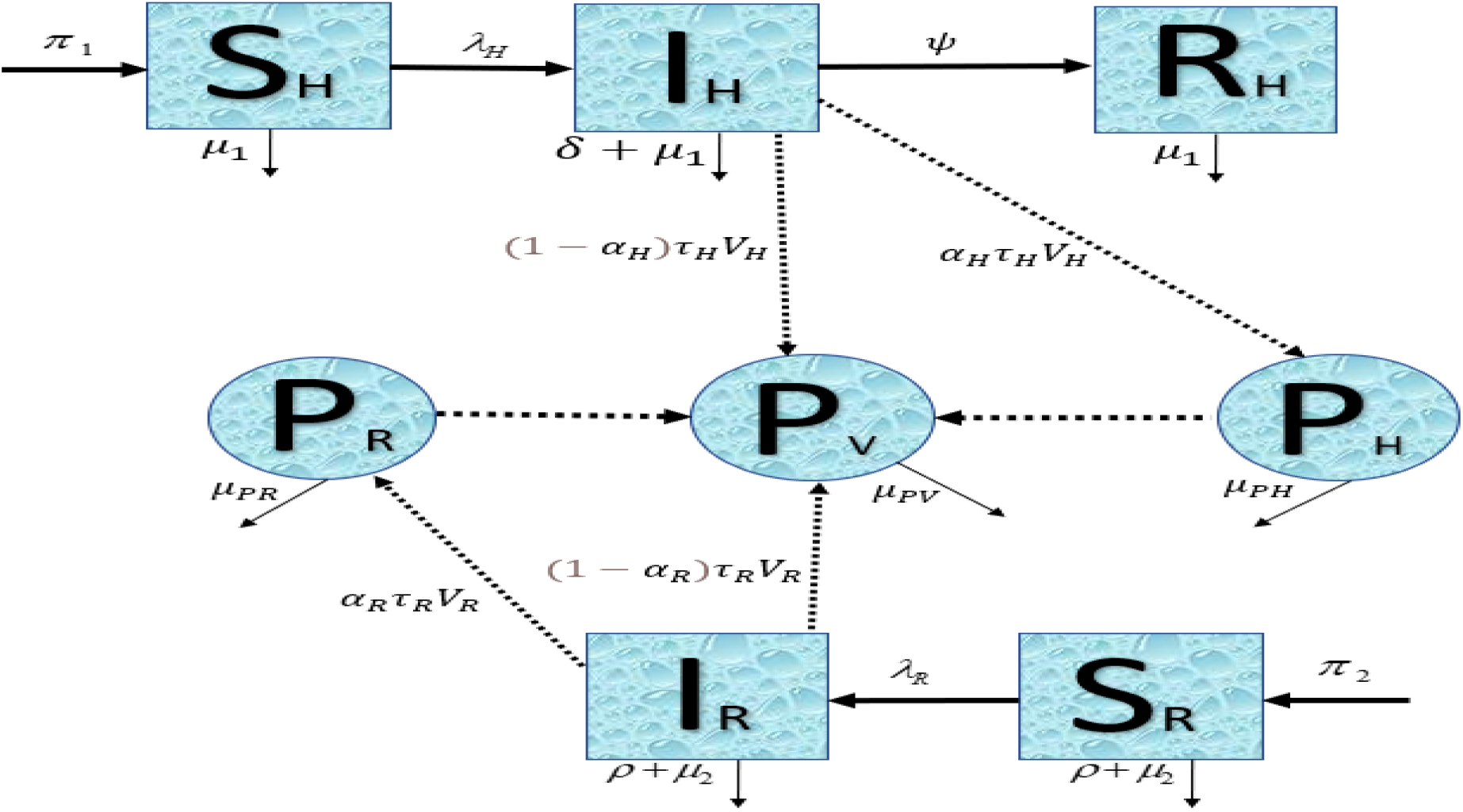
A compartmental representation for the Lassa fever model

**Figure 2:**
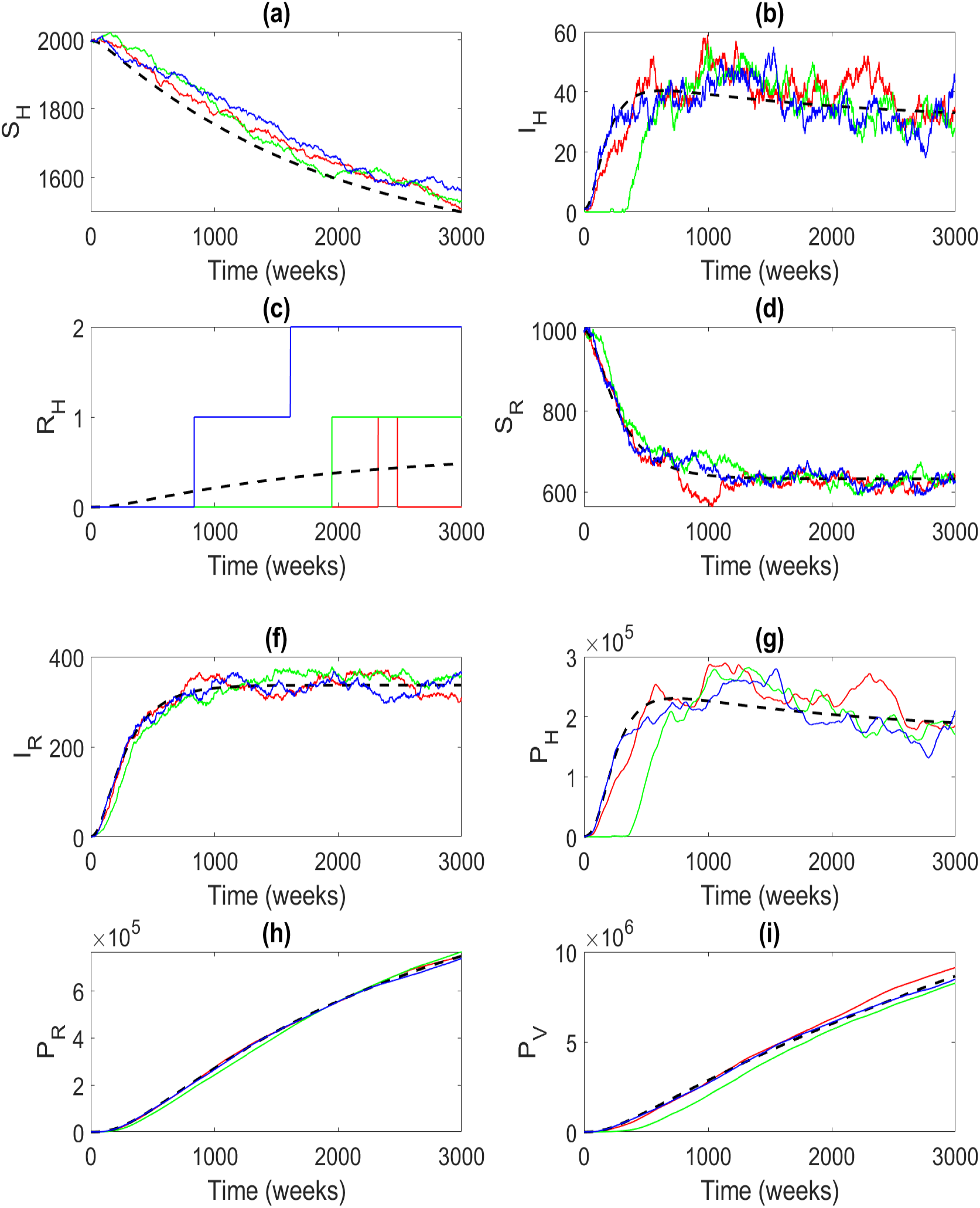
Three sample paths of the CTMC model and the ODE solution (dashed curve) illustrating disease extinction or persistence. Parameter values are as in table 1 and initial conditions are *S*_*H*_ (0) = 1997.8662, *I*_*H*_ (0) = 1, *R*_*H*_ (0) = 0, *S*_*R*_(0) = 1000, *I*_*R*_(0) = 1, *P*_*H*_ (0) = 1, *P*_*R*_(0) = 1, *P*_*V*_ (0) = 1. The probability of extinction is 0.007843158.

### 5.2. Impact of varying transmission parameters on ℙ_0_

To determine the impact of fluctuations in transmission parameter values on ℙ_0_, we vary the parameters by 50% above and below their baseline values as given in table 3. The actual percentage of parameter fluctuations is not easy to establish so we chose 50% arbitrarily just to illustrate the impact of increasing or decreasing transmission parameter values on ℙ_0_. The varied parameters are *β*_*HH*_, *β*_*HR*_, *β*_*HV*_, *β*_*RR*_, *β*_*RV*_, and the results of these fluctuation impacts are shown in tables 4 to 8.

**Table 4:** Probability of disease extinction, ℙ_0_ computed from the branching process. The value of *β*_*HH*_ follows the *±*50% variation of the baseline value with the same initial conditions for the state variables.

| | | | | | -50% ($\beta_{HH} = 0.00875$) | Baseline ($\beta_{HH} = 0.0175$) | +50% ($\beta_{HH} = 0.02625$) |
| --- | --- | --- | --- | --- | --- | --- | --- |
| $x_1$ | $x_2$ | $x_3$ | $x_4$ | $x_5$ | $\mathbb{P}_0$ | $\mathbb{P}_0$ | $\mathbb{P}_0$ |
| 1 | 0 | 0 | 0 | 0 | 0.607023 | 0.605747 | 0.604468 |
| 0 | 1 | 0 | 0 | 0 | 0.223693 | 0.223693 | 0.223693 |
| 0 | 0 | 1 | 0 | 0 | 0.997667 | 0.99533 | 0.992989 |
| 0 | 0 | 0 | 1 | 0 | 0.423947 | 0.423947 | 0.423947 |
| 0 | 0 | 0 | 0 | 1 | 0.137173 | 0.137173 | 0.137138 |
| 1 | 1 | 0 | 0 | 0 | 0.135786796 | 0.135501364 | 0.13521526 |
| 1 | 0 | 1 | 0 | 0 | 0.605606815 | 0.602918162 | 0.600230075 |
| 1 | 0 | 0 | 1 | 0 | 0.25734558 | 0.256804623 | 0.256262395 |
| 1 | 0 | 0 | 0 | 1 | 0.083267166 | 0.083092133 | 0.082895533 |
| 0 | 1 | 1 | 0 | 0 | 0.223171124 | 0.222648354 | 0.222124688 |
| 0 | 1 | 0 | 1 | 0 | 0.094833976 | 0.094833976 | 0.094833976 |
| 0 | 1 | 0 | 0 | 1 | 0.03068464 | 0.03068464 | 0.030676811 |
| 0 | 0 | 1 | 1 | 0 | 0.422957932 | 0.421967168 | 0.420974708 |
| 0 | 0 | 1 | 0 | 1 | 0.136852975 | 0.136532402 | 0.136176525 |
| 0 | 0 | 0 | 1 | 1 | 0.058154082 | 0.058154082 | 0.058139244 |
| 1 | 1 | 1 | 0 | 0 | 0.135470005 | 0.134868572 | 0.134267266 |
| 1 | 1 | 0 | 1 | 0 | 0.057566405 | 0.057445397 | 0.057324104 |
| 1 | 1 | 0 | 0 | 1 | 0.018626282 | 0.018587129 | 0.01854315 |
| 1 | 0 | 1 | 1 | 0 | 0.256745193 | 0.255605346 | 0.25446574 |
| 1 | 0 | 1 | 0 | 1 | 0.083072904 | 0.082704093 | 0.082314352 |
| 1 | 0 | 0 | 1 | 1 | 0.035300865 | 0.035226661 | 0.035143312 |
| 0 | 1 | 1 | 1 | 0 | 0.094612729 | 0.094391102 | 0.094169095 |
| 0 | 1 | 1 | 0 | 1 | 0.030613053 | 0.030541343 | 0.030461736 |
| 0 | 1 | 0 | 1 | 1 | 0.013008661 | 0.013008661 | 0.013005342 |
| 0 | 0 | 1 | 1 | 1 | 0.058018408 | 0.057882502 | 0.057731629 |
| 1 | 1 | 1 | 1 | 0 | 0.057432102 | 0.057177127 | 0.056922205 |
| 1 | 1 | 1 | 0 | 1 | 0.018582827 | 0.018500327 | 0.018413144 |
| 1 | 0 | 1 | 1 | 1 | 0.035218508 | 0.035062152 | 0.034896923 |
| 0 | 1 | 1 | 1 | 1 | 0.012978312 | 0.012947911 | 0.012914161 |
| 1 | 1 | 1 | 1 | 1 | 0.007878134 | 0.007843158 | 0.007806197 |

**Table 5:** Probability of disease extinction, ℙ_0_ computed from the branching process. The value of *β*_*HR*_ follows the *±*50% variation of the baseline value with the same initial conditions for the state variables.

| | | | | | -50% ($\beta_{HR} = 0.00795$) | Baseline ($\beta_{HR} = 0.0159$) | +50% ($\beta_{HR} = 0.02385$) |
| --- | --- | --- | --- | --- | --- | --- | --- |
| $x_1$ | $x_2$ | $x_3$ | $x_4$ | $x_5$ | $\mathbb{P}_0$ | $\mathbb{P}_0$ | $\mathbb{P}_0$ |
| 1 | 0 | 0 | 0 | 0 | 0.605747 | 0.605747 | 0.605746 |
| 0 | 1 | 0 | 0 | 0 | 0.223701 | 0.223693 | 0.223685 |
| 0 | 0 | 1 | 0 | 0 | 0.99533 | 0.99533 | 0.99533 |
| 0 | 0 | 0 | 1 | 0 | 0.423972 | 0.423947 | 0.423923 |
| 0 | 0 | 0 | 0 | 1 | 0.137174 | 0.137173 | 0.137172 |
| 1 | 1 | 0 | 0 | 0 | 0.13550621 | 0.135501364 | 0.135496294 |
| 1 | 0 | 1 | 0 | 0 | 0.602918162 | 0.602918162 | 0.602917166 |
| 1 | 0 | 0 | 1 | 0 | 0.256819767 | 0.256804623 | 0.256789662 |
| 1 | 0 | 0 | 0 | 1 | 0.083092739 | 0.083092133 | 0.08309139 |
| 0 | 1 | 1 | 0 | 0 | 0.222656316 | 0.222648354 | 0.222640391 |
| 0 | 1 | 0 | 1 | 0 | 0.09484296 | 0.094833976 | 0.094825216 |
| 0 | 1 | 0 | 0 | 1 | 0.030685961 | 0.03068464 | 0.030683319 |
| 0 | 0 | 1 | 1 | 0 | 0.421992051 | 0.421967168 | 0.42194328 |
| 0 | 0 | 1 | 0 | 1 | 0.136533397 | 0.136532402 | 0.136531407 |
| 0 | 0 | 0 | 1 | 1 | 0.058157935 | 0.058154082 | 0.058150366 |
| 1 | 1 | 1 | 0 | 0 | 0.134873396 | 0.134868572 | 0.134863526 |
| 1 | 1 | 0 | 1 | 0 | 0.057450839 | 0.057445397 | 0.057439995 |
| 1 | 1 | 0 | 0 | 1 | 0.018587929 | 0.018587129 | 0.018586298 |
| 1 | 0 | 1 | 1 | 0 | 0.255620419 | 0.255605346 | 0.255590454 |
| 1 | 0 | 1 | 0 | 1 | 0.082704696 | 0.082704093 | 0.082703354 |
| 1 | 0 | 0 | 1 | 1 | 0.035228995 | 0.035226661 | 0.035224351 |
| 0 | 1 | 1 | 1 | 0 | 0.094400044 | 0.094391102 | 0.094382382 |
| 0 | 1 | 1 | 0 | 1 | 0.030542658 | 0.030541343 | 0.030540028 |
| 0 | 1 | 0 | 1 | 1 | 0.013009988 | 0.013008661 | 0.013007365 |
| 0 | 0 | 1 | 1 | 1 | 0.057886338 | 0.057882502 | 0.057878804 |
| 1 | 1 | 1 | 1 | 0 | 0.057182543 | 0.057177127 | 0.057171751 |
| 1 | 1 | 1 | 0 | 1 | 0.018501123 | 0.018500327 | 0.0184995 |
| 1 | 0 | 1 | 1 | 1 | 0.035064475 | 0.035062152 | 0.035059854 |
| 0 | 1 | 1 | 1 | 1 | 0.012949232 | 0.012947911 | 0.01294662 |
| 1 | 1 | 1 | 1 | 1 | 0.007843958 | 0.007843158 | 0.007842363 |

**Table 6:**
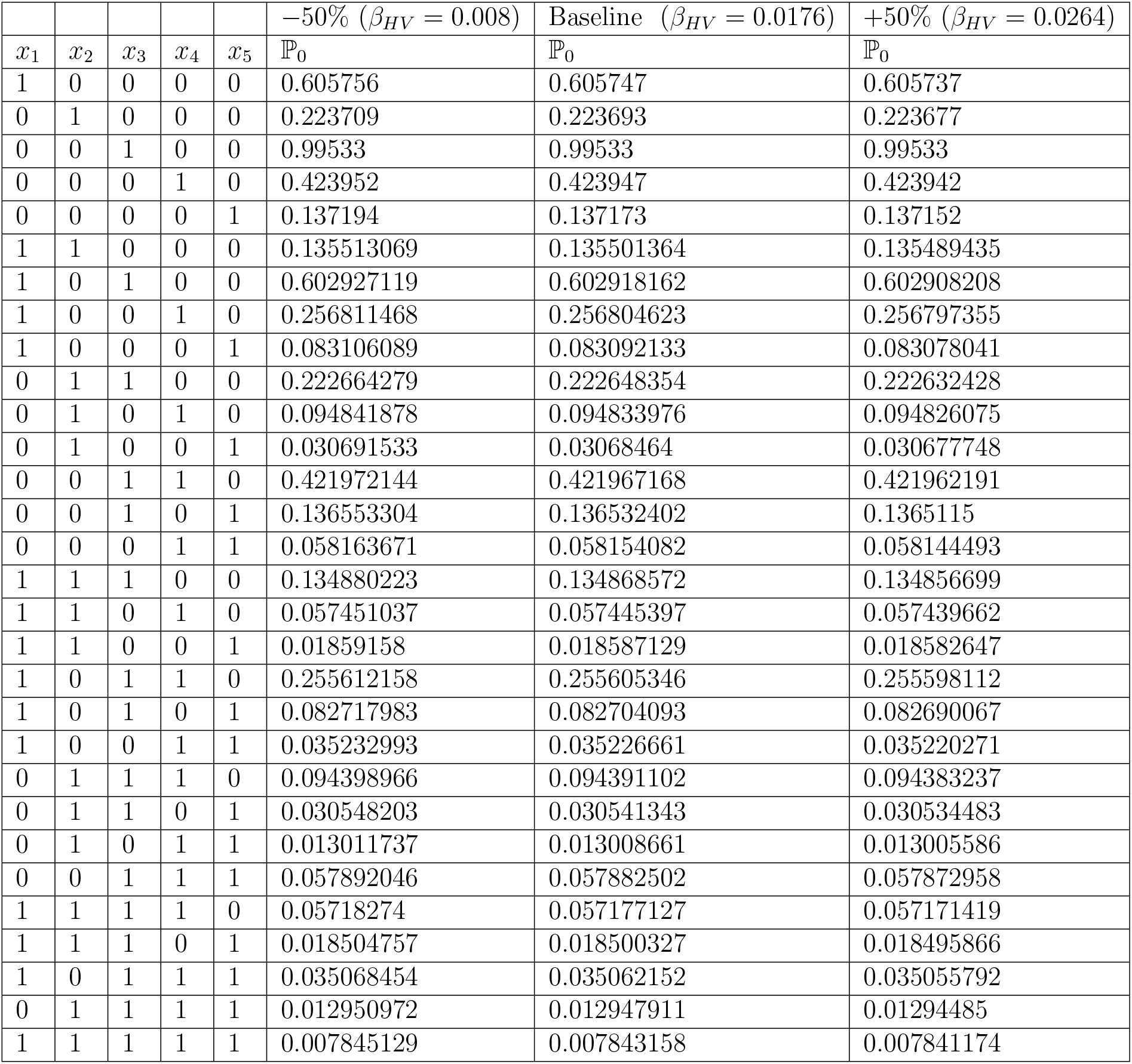
Probability of disease extinction, ℙ_0_ computed from the branching process. The value of *β*_*HV*_ follows the *±*50% variation of the baseline value with the same initial conditions for the state variables.

| | | | | | $-50\%$ ($\beta_{HV} = 0.008$) | Baseline ($\beta_{HV} = 0.0176$) | $+50\%$ ($\beta_{HV} = 0.0264$) |
| --- | --- | --- | --- | --- | --- | --- | --- |
| $x_1$ | $x_2$ | $x_3$ | $x_4$ | $x_5$ | $\mathbb{P}_0$ | $\mathbb{P}_0$ | $\mathbb{P}_0$ |
| 1 | 0 | 0 | 0 | 0 | 0.605756 | 0.605747 | 0.605737 |
| 0 | 1 | 0 | 0 | 0 | 0.223709 | 0.223693 | 0.223677 |
| 0 | 0 | 1 | 0 | 0 | 0.99533 | 0.99533 | 0.99533 |
| 0 | 0 | 0 | 1 | 0 | 0.423952 | 0.423947 | 0.423942 |
| 0 | 0 | 0 | 0 | 1 | 0.137194 | 0.137173 | 0.137152 |
| 1 | 1 | 0 | 0 | 0 | 0.135513069 | 0.135501364 | 0.135489435 |
| 1 | 0 | 1 | 0 | 0 | 0.602927119 | 0.602918162 | 0.602908208 |
| 1 | 0 | 0 | 1 | 0 | 0.256811468 | 0.256804623 | 0.256797355 |
| 1 | 0 | 0 | 0 | 1 | 0.083106089 | 0.083092133 | 0.083078041 |
| 0 | 1 | 1 | 0 | 0 | 0.222664279 | 0.222648354 | 0.222632428 |
| 0 | 1 | 0 | 1 | 0 | 0.094841878 | 0.094833976 | 0.094826075 |
| 0 | 1 | 0 | 0 | 1 | 0.030691533 | 0.03068464 | 0.030677748 |
| 0 | 0 | 1 | 1 | 0 | 0.421972144 | 0.421967168 | 0.421962191 |
| 0 | 0 | 1 | 0 | 1 | 0.136553304 | 0.136532402 | 0.1365115 |
| 0 | 0 | 0 | 1 | 1 | 0.058163671 | 0.058154082 | 0.058144493 |
| 1 | 1 | 1 | 0 | 0 | 0.134880223 | 0.134868572 | 0.134856699 |
| 1 | 1 | 0 | 1 | 0 | 0.057451037 | 0.057445397 | 0.057439662 |
| 1 | 1 | 0 | 0 | 1 | 0.01859158 | 0.018587129 | 0.018582647 |
| 1 | 0 | 1 | 1 | 0 | 0.255612158 | 0.255605346 | 0.255598112 |
| 1 | 0 | 1 | 0 | 1 | 0.082717983 | 0.082704093 | 0.082690067 |
| 1 | 0 | 0 | 1 | 1 | 0.035232993 | 0.035226661 | 0.035220271 |
| 0 | 1 | 1 | 1 | 0 | 0.094398966 | 0.094391102 | 0.094383237 |
| 0 | 1 | 1 | 0 | 1 | 0.030548203 | 0.030541343 | 0.030534483 |
| 0 | 1 | 0 | 1 | 1 | 0.013011737 | 0.013008661 | 0.013005586 |
| 0 | 0 | 1 | 1 | 1 | 0.057892046 | 0.057882502 | 0.057872958 |
| 1 | 1 | 1 | 1 | 0 | 0.05718274 | 0.057177127 | 0.057171419 |
| 1 | 1 | 1 | 0 | 1 | 0.018504757 | 0.018500327 | 0.018495866 |
| 1 | 0 | 1 | 1 | 1 | 0.035068454 | 0.035062152 | 0.035055792 |
| 0 | 1 | 1 | 1 | 1 | 0.012950972 | 0.012947911 | 0.01294485 |
| 1 | 1 | 1 | 1 | 1 | 0.007845129 | 0.007843158 | 0.007841174 |

**Table 7:** Probability of disease extinction, ℙ_0_ computed from the branching process. The value of *β*_*RR*_ follows the *±*50% variation of the baseline value with the same initial conditions for the state variables.

| | | | | | $-50\%$ ($\beta_{RR} = 0.0061$) | Baseline ($\beta_{RR} = 0.0122$) | $+50\%$ ($\beta_{RR} = 0.0183$) |
| --- | --- | --- | --- | --- | --- | --- | --- |
| $x_1$ | $x_2$ | $x_3$ | $x_4$ | $x_5$ | $\mathbb{P}_0$ | $\mathbb{P}_0$ | $\mathbb{P}_0$ |
| 1 | 0 | 0 | 0 | 0 | 0.61064 | 0.605747 | 0.603408 |
| 0 | 1 | 0 | 0 | 0 | 0.288866 | 0.223693 | 0.188805 |
| 0 | 0 | 1 | 0 | 0 | 0.995388 | 0.99533 | 0.995303 |
| 0 | 0 | 0 | 1 | 0 | 0.616338 | 0.423947 | 0.319526 |
| 0 | 0 | 0 | 0 | 1 | 0.147882 | 0.137173 | 0.132054 |
| 1 | 1 | 0 | 0 | 0 | 0.176393134 | 0.135501364 | 0.113926447 |
| 1 | 0 | 1 | 0 | 0 | 0.607823728 | 0.602918162 | 0.600573793 |
| 1 | 0 | 0 | 1 | 0 | 0.376360636 | 0.256804623 | 0.192804545 |
| 1 | 0 | 0 | 0 | 1 | 0.090302664 | 0.083092133 | 0.07968244 |
| 0 | 1 | 1 | 0 | 0 | 0.28753375 | 0.222648354 | 0.187918183 |
| 0 | 1 | 0 | 1 | 0 | 0.178039093 | 0.094833976 | 0.060328106 |
| 0 | 1 | 0 | 0 | 1 | 0.042718082 | 0.03068464 | 0.024932455 |
| 0 | 0 | 1 | 1 | 0 | 0.613495449 | 0.421967168 | 0.318025186 |
| 0 | 0 | 1 | 0 | 1 | 0.147199968 | 0.136532402 | 0.131433742 |
| 0 | 0 | 0 | 1 | 1 | 0.091145296 | 0.058154082 | 0.042194686 |
| 1 | 1 | 1 | 0 | 0 | 0.175579609 | 0.134868572 | 0.113391335 |
| 1 | 1 | 0 | 1 | 0 | 0.108717792 | 0.057445397 | 0.036402462 |
| 1 | 1 | 0 | 0 | 1 | 0.026085369 | 0.018587129 | 0.015044443 |
| 1 | 0 | 1 | 1 | 0 | 0.374624861 | 0.255605346 | 0.191898942 |
| 1 | 0 | 1 | 0 | 1 | 0.089886189 | 0.082704093 | 0.079308172 |
| 1 | 0 | 0 | 1 | 1 | 0.055656964 | 0.035226661 | 0.025460611 |
| 0 | 1 | 1 | 1 | 0 | 0.177217976 | 0.094391102 | 0.060044745 |
| 0 | 1 | 1 | 0 | 1 | 0.042521066 | 0.030541343 | 0.024815348 |
| 0 | 1 | 0 | 1 | 1 | 0.026328777 | 0.013008661 | 0.007966568 |
| 0 | 0 | 1 | 1 | 1 | 0.090724934 | 0.057882502 | 0.041996498 |
| 1 | 1 | 1 | 1 | 0 | 0.108216385 | 0.057177127 | 0.03623148 |
| 1 | 1 | 1 | 0 | 1 | 0.025965064 | 0.018500327 | 0.014973779 |
| 1 | 0 | 1 | 1 | 1 | 0.055400274 | 0.035062152 | 0.025341023 |
| 0 | 1 | 1 | 1 | 1 | 0.026207349 | 0.012947911 | 0.007929149 |
| 1 | 1 | 1 | 1 | 1 | 0.016003255 | 0.007843158 | 0.004784512 |

**Table 8:** Probability of disease extinction, ℙ_0_ computed from the branching process. The value of *β*_*RV*_ follows the *±*50% variation of the baseline value with the same initial conditions for the state variables.

| | | | | | $-50\%$ ($\beta_{RV} = 0.00615$) | Baseline ($\beta_{RV} = 0.0123$) | $+50\%$ ($\beta_{RV} = 0.01845$) |
| --- | --- | --- | --- | --- | --- | --- | --- |
| $x_1$ | $x_2$ | $x_3$ | $x_4$ | $x_5$ | $\mathbb{P}_0$ | $\mathbb{P}_0$ | $\mathbb{P}_0$ |
| 1 | 0 | 0 | 0 | 0 | 0.665456 | 0.605747 | 0.585159 |
| 0 | 1 | 0 | 0 | 0 | 0.325497 | 0.223693 | 0.18898 |
| 0 | 0 | 1 | 0 | 0 | 0.996035 | 0.99533 | 0.995088 |
| 0 | 0 | 0 | 1 | 0 | 0.458592 | 0.423947 | 0.413301 |
| 0 | 0 | 0 | 0 | 1 | 0.267852 | 0.137173 | 0.0921153 |
| 1 | 1 | 0 | 0 | 0 | 0.216603932 | 0.135501364 | 0.110583348 |
| 1 | 0 | 1 | 0 | 0 | 0.662817467 | 0.602918162 | 0.582284699 |
| 1 | 0 | 0 | 1 | 0 | 0.305172798 | 0.256804623 | 0.2418468 |
| 1 | 0 | 0 | 0 | 1 | 0.178243721 | 0.083092133 | 0.053902097 |
| 0 | 1 | 1 | 0 | 0 | 0.324206404 | 0.222648354 | 0.18805173 |
| 0 | 1 | 0 | 1 | 0 | 0.14927032 | 0.094833976 | 0.078105623 |
| 0 | 1 | 0 | 0 | 1 | 0.087185022 | 0.03068464 | 0.017407949 |
| 0 | 0 | 1 | 1 | 0 | 0.456773683 | 0.421967168 | 0.411270865 |
| 0 | 0 | 1 | 0 | 1 | 0.266789967 | 0.136532402 | 0.09166283 |
| 0 | 0 | 0 | 1 | 1 | 0.122834784 | 0.058154082 | 0.038071346 |
| 1 | 1 | 1 | 0 | 0 | 0.215745097 | 0.134868572 | 0.110040162 |
| 1 | 1 | 0 | 1 | 0 | 0.09933283 | 0.057445397 | 0.045704208 |
| 1 | 1 | 0 | 0 | 1 | 0.058017796 | 0.018587129 | 0.010186418 |
| 1 | 0 | 1 | 1 | 0 | 0.303962788 | 0.255605346 | 0.240658848 |
| 1 | 0 | 1 | 0 | 1 | 0.177536984 | 0.082704093 | 0.05363733 |
| 1 | 0 | 0 | 1 | 1 | 0.081741144 | 0.035226661 | 0.022277791 |
| 0 | 1 | 1 | 1 | 0 | 0.148678463 | 0.094391102 | 0.077721968 |
| 0 | 1 | 1 | 0 | 1 | 0.086839334 | 0.030541343 | 0.017322442 |
| 0 | 1 | 0 | 1 | 1 | 0.039982354 | 0.013008661 | 0.007194723 |
| 0 | 0 | 1 | 1 | 1 | 0.122347744 | 0.057882502 | 0.037884339 |
| 1 | 1 | 1 | 1 | 0 | 0.098938976 | 0.057177127 | 0.045479709 |
| 1 | 1 | 1 | 0 | 1 | 0.057787756 | 0.018500327 | 0.010136383 |
| 1 | 0 | 1 | 1 | 1 | 0.081417041 | 0.035062152 | 0.022168362 |
| 0 | 1 | 1 | 1 | 1 | 0.039823824 | 0.012947911 | 0.007159382 |
| 1 | 1 | 1 | 1 | 1 | 0.026501002 | 0.007843158 | 0.004189377 |

Table 3 shows the different probabilities of disease extinction associated with different combinations of initial contributors to infection. We see from table 4 to 6 that variations in the contact rate between susceptible humans and HCPL, contact rate between susceptible humans and RCPL, and contact rate between susceptible humans and VCPL is associated with the least variation in the probability of extinction. This is followed by a variation in the contact rate between susceptible rodents and the RCPL as seen in table 7. A smaller value of *β*_*RR*_ is associated with a greater probabilty of disease extinction. Also, from table 8, we see that variations in *β*_*RV*_, the contact rate between susceptible rodents and the virus community pathogen load is associated with the greatest variation in the probability of extinction. When we compare results from table 3 with tables 4 to 8, we see that an increase in the transmission rates from the rodents and environment population leads to a lower probability of extinction. This underscores the potential for disease transmission arising from both environmental factors and the presence of rodents.

Strategies aimed at mitigating the public health impact of this virus should prioritize reducing indirect human-rodent contact through measures such as improved sanitation and fumigation processes while intervention measures on other contact routes should not be neglected. These interventions have the potential to lower the probability of human infection by minimizing the interaction between humans and rodents in contaminated environments.

## 6. Summary of Results

Addressing the rising incidence of LASV in endemic regions poses a significant challenge in the broader battle against LF. To contribute to this ongoing effort, we developed and scrutinized a CTMC model capturing the dynamics of LF within human, rodent, and virus populations, building upon the deterministic model (5). The objectives of this investigation were to explore the influence of demographic stochasticity on the persistence of LASV within these populations, and assess the impact of varying transmission rates on the probability of disease extinction. We conducted an analysis of the existence and stability of equilibria in the deterministic model. The results demonstrated that steady states exist and are locally asymptotically stable when *R*_0_ *>* 1. Numerical simulations confirmed the presence of the virus when the reproduction number surpassed unity. We utilized a multitype branching process to examine the nonlinear CTMC model in the vicinity of the DFE. This analysis yielded an expression that was employed to quantify the probability of disease extinction and persistence. The probabilities were governed by the extinction threshold *ρ*(***M***).

Our results show an increase in the disease extinction when LASV is introduced at the beginning of an epidemic, whether through HCPL or an infected human in combination with RCPL and infected rodent. This probability diminishes when LASV is introduced through VCPL in combination with other transmission routes. This observation aligns with the notion that the presence of multiple infection routes amplifies the probability of disease persistence, highlighting the complex interplay of transmission dynamics. Our study also highlights the impact of variations in contact rates on viral transmission and, consequently, on the probability of disease extinction. We observed that alterations in the contact rate between susceptible rodents and the VCPL exhibited the highest sensitivity, underscoring its pivotal role in influencing disease dynamics. Nevertheless, perturbations in other contact rates were also found to induce changes in the probability of disease extinction, emphasizing the interconnected nature of these transmission pathways. These insights underscore the intricate relationship between the introduced pathways and contact rates, shedding light on the factors that govern the persistence or extinction of LASV within a population. Such knowledge is crucial for developing targeted and effective strategies in the management and control of Lassa fever.

## Data Availability

All data used are available online at https://www.ncdc.gov.ng/diseases/sitreps/

## Appendix A. Sample Appendix Section

This is the Appendix.

